# A Structural Heart-Brain Axis Mediates the Association Between Cardiovascular Risk and Cognitive Function

**DOI:** 10.1101/2022.09.15.22279275

**Authors:** Akshay Jaggi, Eleanor L.S. Conole, Zahra Raisi-Estabragh, Polyxeni Gkontra, Celeste McCracken, Stefan Neubauer, Steffen E. Petersen, Simon Cox, Karim Lekadir

**Affiliations:** Facultat de Matemàtiques i Informàtica, Universitat de Barcelona, Barcelona; Lothian Birth Cohorts, Department of Psychology, The University of Edinburgh, 7 George Square, Edinburgh EH8 9JZ, UK; William Harvey Research Institute, NIHR Barts Biomedical Research Centre, Queen Mary University of London, Charterhouse Square, London, EC1M 6BQ, UK; Barts Heart Centre, St Bartholomew’s Hospital, Barts Health NHS Trust, West Smithfield, EC1A 7BE, London, UK; Division of Cardiovascular Medicine, Radcliffe Department of Medicine, University of Oxford, National Institute for Health Research Oxford Biomedical Research Centre, Oxford University Hospitals NHS Foundation Trust, Oxford, OX3 9DU, UK; Health Data Research UK, London, UK; Alan Turing Institute, London, UK

## Abstract

Elevated vascular disease risk associates with poorer cognitive function, but the mechanism for this link is poorly understood. A leading theory, the structural-functional model argues that vascular risk may drive adverse cardiac remodelling, which in turn leads to chronic cerebral hypoperfusion and subsequent brain structural damage. This model predicts that variation in heart and brain structure should associate with both greater vascular risk and lower cognitive function. This study tests that prediction in a large sample of the UK Biobank (N=11,962). We assemble and summarise vascular risk factors, cardiac magnetic resonance radiomics, brain structural and diffusion MRI indices, and cognitive assessment. We also extract ‘heart-brain axes’ capturing the covariation in heart and brain structure. Many heart and brain measures partially explain the vascular risk – cognitive function association, like left ventricular end-diastolic volume and grey matter volume. Notably, a heart-brain axis, capturing correlation between lower myocardial intensity, lower grey matter volume, and poorer thalamic white matter integrity, completely mediates the association, supporting the structural-functional model. Our findings also complicate this theory by finding that brain structural variation cannot completely explain the heart structure – cognitive function association. Our results broadly offer evidence for the structural functional hypothesis, identify imaging biomarkers for this association by considering covariation in heart and brain structure, and generate novel hypotheses about how cardiovascular risk may link to cognitive function.

## Introduction

With ageing populations throughout the world, cognitive decline now affects an increasingly large portion of society and contributes to significant financial burden and death.^1,2^ Of the drivers of age-related cognitive decline, neurovascular health has gained attention due to its widespread impact and relative ease of intervention.^3–6^

Substantial work has shown diverse associations between vascular disease risk factors (VRFs, such as diabetes, high body mass index (BMI), and hypertension) and cognitive function (CF). Greater vascular risk in middle and old age associates with both poorer cognitive function and accelerated cognitive decline,^7–11^ and controlling vascular risk factors can lead to a decrease in onset of mild cognitive impairment.^12^

Better understanding of the mechanism of this heart-brain axis will facilitate biomarker development and treatment discovery for neurovascular health. A few mechanistic theories exist but lack evidence. One popular model, the structural-functional model, argues that VRFs might drive pathologic cardiac and cerebrovascular remodelling, which could then result in chronic cerebral hypoperfusion, brain structural damage, and poorer CF.^6,13–16^ Direct evidence for this theory has remained unclear but could be found by simultaneously measuring vascular risk factors, cognitive function, and heart and brain structure.

Cardiac and brain imaging derived phenotypes (IDPs) have become popular methods for measuring heart and brain structure due to their minimally invasive nature and widespread use. Both are strong candidate biomarkers of the modest but well-replicated association between elevated vascular risk and lower cognitive function in middle and older age.^7,17^ However, to-date, most of our knowledge about associations between 1) VRFs and 2) cardiac structure, 3) brain structure and 4) cognitive measures come from separate reports, which only simultaneously consider two phenotypes of interest.^7,18–21^ Several recent works have indicated the value in extending analyses across three of the four phenotype categories above; for example, lower grey matter volume can explain part of the association between hypertension, greater BMI, and lower performance on some UK Biobank cognitive exams.^17,22–24^ However, these studies have only studied a restricted set of risk factors or neuroimaging measures, and have yet to incorporate heart structure and model the heart-brain axis in this context.

We hypothesise that, for the structural-functional model to adequately explain the VRF-CF association, separate heart and brain structures should associate with both greater vascular risk and lower cognitive function. In other words, heart and brain structural variation should mediate the VRF-CF association. Additionally, heart mediators should associate with brain mediators. Finally, for all steps of the structural-functional model to be supported by the data, heart structural variation should mediate the VRF - brain structure association, and brain structural variation should mediate the heart structure - CF association. The extent to which these associations all align in a cohort of subjects modelled together is understudied.^17,24,25^ Furthermore, the relative strength of the association between cardiac and brain structural features and the disease endpoints (vascular risk and cognitive decline) is unknown. Along with validating the structural functional hypothesis, this comparative approach could identify novel biomarkers associated specifically with the VRF-CF association (rather than each dataset alone) and guide future decision-making comparing and prioritising organ-specific interventions in vascular and cognitive health.^2,4,25^

To test the structural-functional hypothesis, in this work, we measure the extent that variation in heart and brain structure explains the association between vascular risk and cognitive function in the UK Biobank. We gather vascular risk factors, cognitive exam performance, CMR radiomics features, and brain MRI IDPs for 11,962 UK Biobank participants. We perform dimensionality reduction on all datasets separately. We discover novel measures of the heart-brain axis by capturing correlated variance in heart and brain imaging. We compute single and multiple mediation models asking how well imaging latent variables explain the VRF - CF association.

We then measure how well imaging latent variables explain associations between individual VRFs and cognitive exams. We finally explore how well individual heart and brain structural measures mediate the VRF - CF association. Along with myriad smaller mediating effects, we find that myocardial intensity, grey matter volume, and thalamic white matter tract integrity all associate with each other, and a joint factor capturing their variability most strongly associates with both elevated vascular risk and poorer cognitive function.

## Results

### Quantifying Heart Brain Axes

After our data preparation pipeline yielded 11,962 subjects (**Supplementary Figure 1, Supplementary Table 1**), we quantified key axes of variation in all four of our datasets. We extracted latent measures of vascular risk (gVRF), cognitive function (g), and brain structure as reported previously (see **Methods**).^7,17,19,26,27^ Along with traditional measures, we performed PCA of heart and brain imaging separately and a novel CCA to capture correlated variability in heart and brain structure (**Figure 1**). For cardiac radiomics, the first three PCs explain 25, 20, and 12% of the variance and represent myocardial size, intensity, and textural complexity respectively (**Supplementary Figure 4, Supplementary Table 4**). For brain MRI indices, the first three PCs explain 30, 12, and 8% of the variance and represent WM integrity of the fasciculi and thalamic radiata, WM integrity of the corticospinal tract, and brain volume respectively (**Supplementary Figure 6, Supplementary Table 6**). For the joint heart brain axes, the first three modes have a Pearson correlation of 0.71, 0.48, and 0.32 respectively (**Supplementary Figure 7, Supplementary Table 7**). Based on the loadings, we interpreted that the heart brain axes correspond to 1) heart and brain volume, 2) end-systolic myocardial intensity, grey matter and thalamic volume, and thalamic radiation WM integrity, and 3) end-diastolic myocardial intensity and WM pathology.

**Figure 1:**
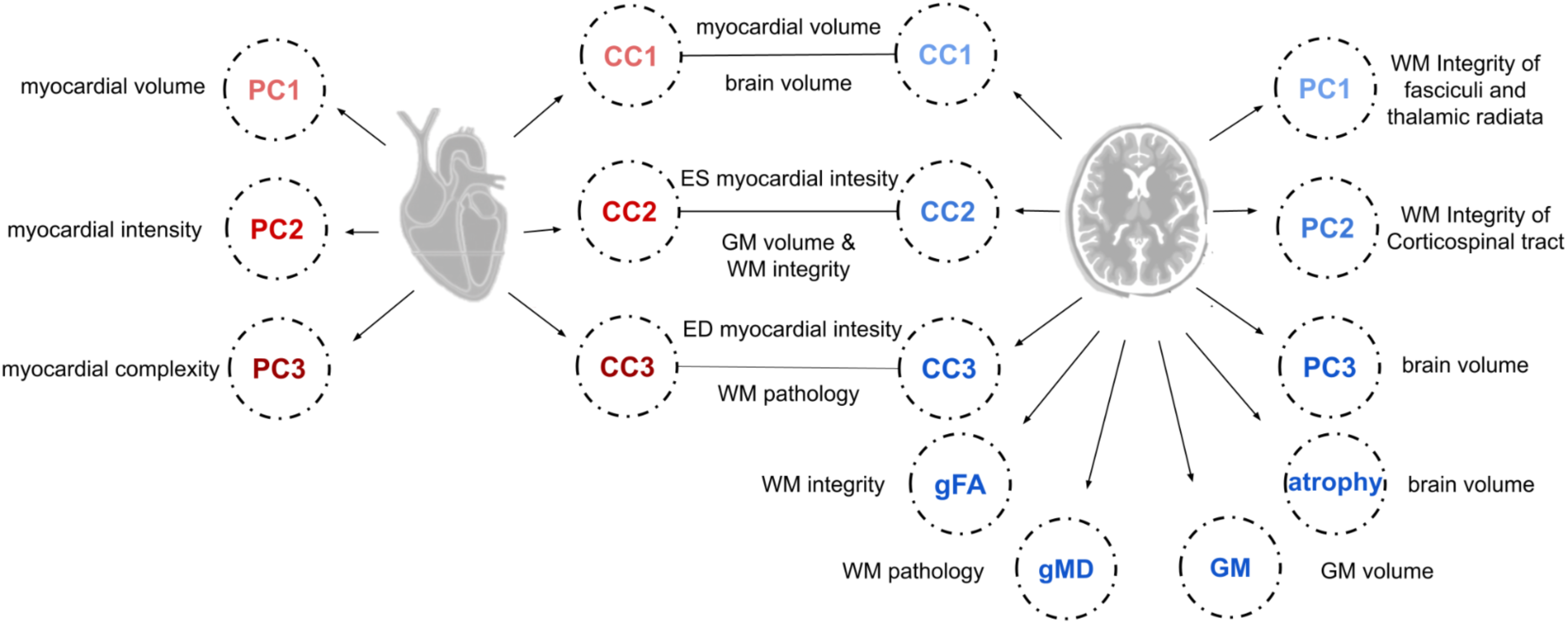
Latent Factors

### Descriptive Statistics

Nearly all latent variables have a significant association with age and sex (**Supplementary Figure 8, Supplementary Tables 8, 9**). Older subjects show lower aggregate performance on cognitive exams (β=-0.183) and greater vascular risk (β=0.171).^7,19^ Among the heart structural latents, old age associates with slightly greater myocardial volume (CMR PC1, β=0.035), lower myocardial intensity (PC2, β=-0.173), and lower myocardial textural complexity (PC3, β=-0.109).^28^

A schematic illustrating all of the extracted latent factors and a simple interpretation of their meaning. The loadings for all the factors can be found in the **Supplementary Tables** and more detailed interpretations of the meaning of each factor can be found in the **Supplementary Methods**.

Among the brain structural latents, old age associates with lower total and grey matter volume and lower white matter integrity (β range -0.363 to -0.249). Age also strongly negatively associates with the components of the second CCA mode, representing lower myocardial intensity, grey matter and thalamic volume, and thalamic white matter integrity (β range -0.591 to -0.441).

### Associations Between Vascular Risk, Heart, Brain, and Cognition

Associations among each pair of latent variables were modeled separately, controlling for age and sex (**Figure 2, Supplementary Tables 10, 11**). There is a small but significant negative association between gVRF and g (β=-0.036), consistent with prior reports.^7,17^ Many imaging latents across heart and brain associate with both greater gVRF and lower g: lower myocardial intensity, lower total and grey matter volume, and lower white matter tract integrity (**Figure 2, Supplementary Table 11**).

**Figure 2:**
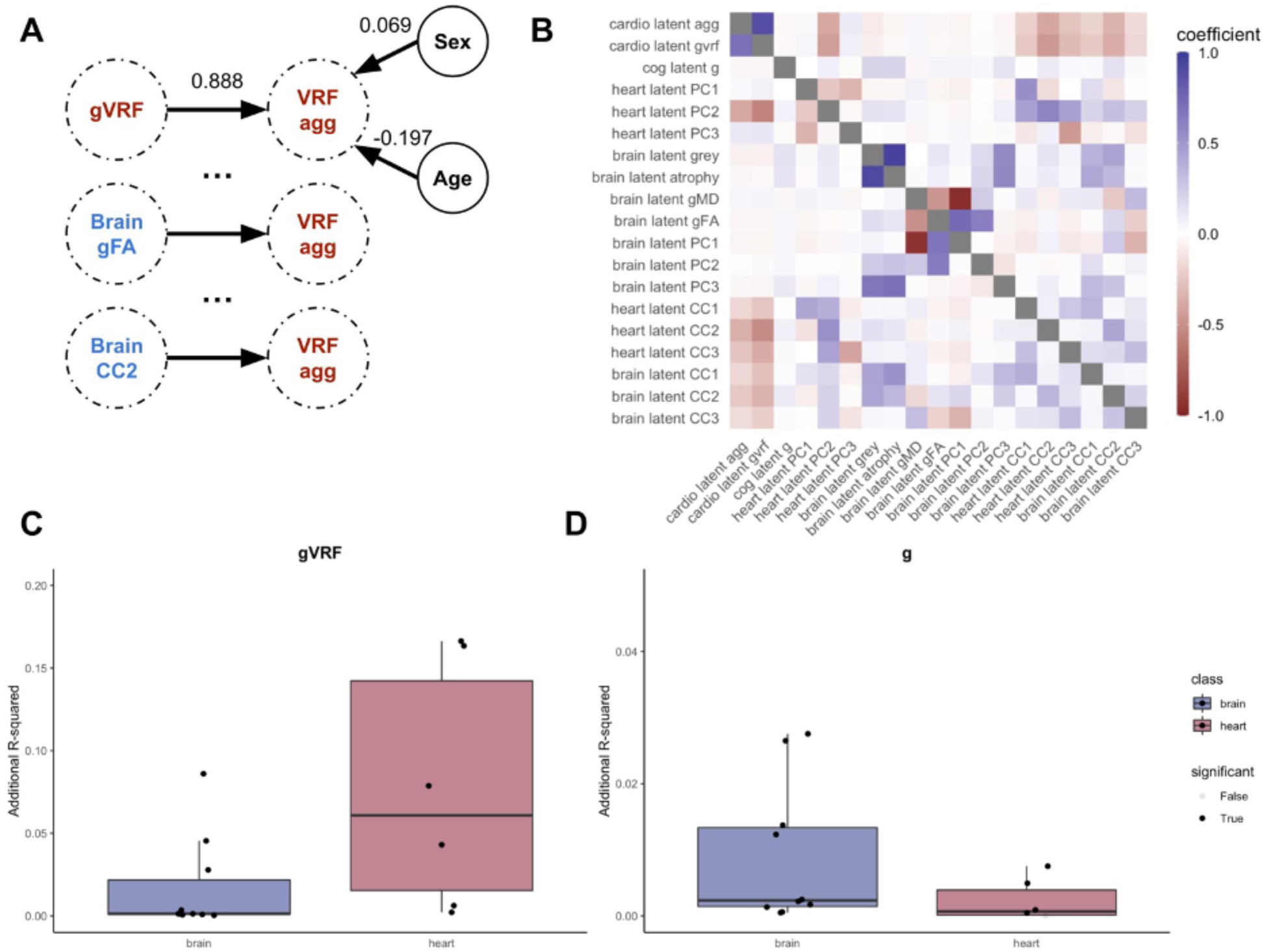
Pairwise Latent Associations

We modelled the association between every pair of latent variables. (A) A schematic diagram of the modelling process. Every latent variable (e.g.VRF agg) is linearly modelled as a function of another latent variable (e.g. gVRF), sex, and age. The derived coefficients for the example first model are illustrated. We repeat this for every variable, and the coefficient from these analyses compose the first row of the adjacent heatmap. (B) Heatmap of standardised coefficients from all 342 separate pairwise linear models. Each row lists the dependent variable, and each column lists the independent variable in the linear models. (C) With gVRF set as the dependent variable, we compare the R-squared of the linear model for each latent grouped by whether it was derived from the heart or brain imaging. (D) With g set as the dependent variable, we compare the R-squared of the linear model for each latent grouped by whether it was derived from the heart or brain imaging. All model estimates reported in **Supplementary Table 11**.

All of the heart PCs explained at least an order of magnitude more variance in gVRF (additional R^2^: 0.002–0.166) than in g (aR^2^: 0–0.004) (**Figure 2**). Similarly, the brain volume latents (atrophy, grey matter volume, PC3) explained at least an order of magnitude greater variance in g (aR^2^: 0.012–0.027) than in gVRF (aR^2^: 0.0006–0.003). Interestingly, the second joint factor (CC2) explains more similar amounts of variance in both g (aR^2^: 0.009–0.013) and gVRF (aR^2^: 0.089–0.164), and it explains at least an order of magnitude more variance in both g and gVRF than the white matter latents. This suggests that leveraging information from both heart and brain structure is useful in deriving factors that explain a relatively large and equal amount of variance in both vascular risk and cognitive function.

### Matched Analysis

Aware that the latent measures are all in arbitrary units, we used propensity score matching to provide more practically interpretable information on how those with high and low vascular risk differ across heart, brain and cognitive measures, in native units. We assembled two groups of 425 subjects matched by sex, age, head size, and BSA (**Supplementary Table 12**). On average, when compared to matched individuals with no VRFs, subjects with 4 or more VRFs have 13.09 mL (8.29%) lower LVEDV, 7.56 mL (11.50%) lower LVESV, and 5.52 mL (5.99%) lower LVSV. Consistent with mild ventricular hypertrophy, the subjects with 4 or more VRFs have 1.51% (2.58%) greater ejection fraction. We find lower average intensities of the myocardium in end-systole (23.53%) and diastole (19.65%). We also find greater uniformity of the myocardial tissue appearance (5.25–8.37%). These subjects also have 14,357 mm^3^ (2.31%) less grey matter volume and additionally lower subcortical volumes. They also have lower FA in many tracts (range 0.96% and 1.92%). Compared to matched healthy controls, subjects with 4 or more VRFs also score on average 0.48 (6.67%) fewer points on verbal-numerical reasoning. These subjects also have notable differences in their latent measures, like greater myocardial size, poorer white matter tracts, and lower second heart-brain axis (myocardial intensity, grey matter volume, thalamic WM tract integrity). Simply summing risk factors correlates with gVRF (**Figure 2, Supplementary Table 11**), and this matched analysis shows that the sum manifests with clinically observable phenotypes in heart imaging, brain imaging, and cognitive exam performance.

### Latent Single Mediation Modelling

Initially, we asked the degree to which each brain or heart measure, in isolation, mediates the association between vascular risk and CF. Results are presented in **Figure 3, Supplementary Tables 13, 14**. Consistent with prior reports, measures of brain structure - irrespective of how they were measured - only modestly mediated the association (4.97–38.12%), with white matter measures being the smallest, but still significant, mediators. However, myocardial intensity (heart PC2) and the heart-brain axis capturing myocardial intensity, grey matter volume, and thalamic white matter integrity (CC2) all completely mediate the gVRF-g association (117%-150%; attenuated to be indistinguishable from β =0 in each case). For example, one standard deviation (SD) lower gVRF associates with 0.55 standard deviation lower latent myocardial intensity. This 0.55 SD lower intensity associates with 0.043 SD lower cognitive function.

**Figure 3:**
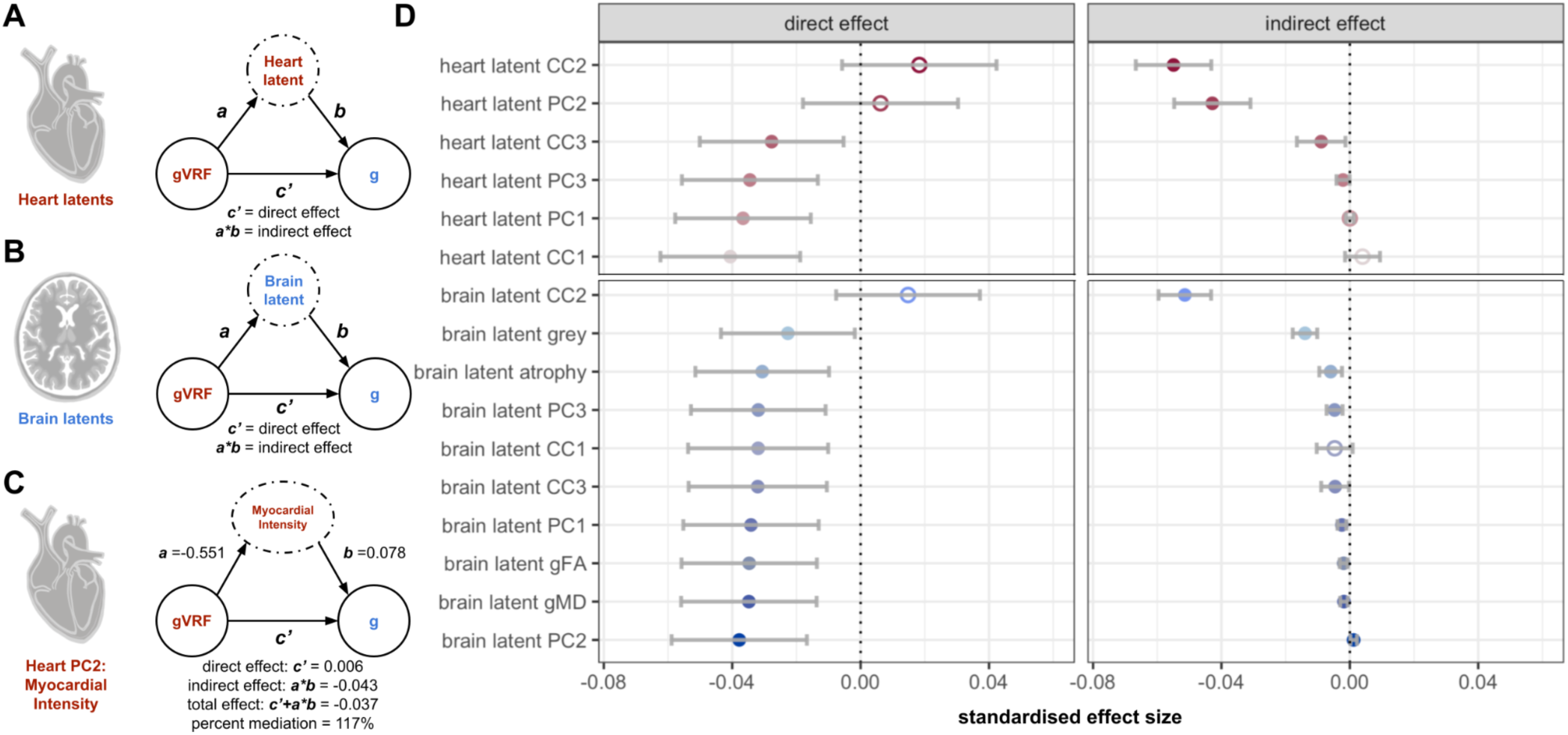
Latent Single Mediation Modelling

As a control, we address two possible counterarguments: (1) that the BMI - cognitive function association is the only VRF well explained by myocardial intensity and (2) that myocardial intensity is just a proxy for myocardial size. First, since gVRF most strongly weights BMI and WHR (**Supplementary Table 2**), it’s possible that the gVRF-g association is driven primarily by BMI and that myocardial intensity only mediates the BMI - g association. However, covarying for BMI partly attenuated, but did not remove, myocardial intensity’s mediation of the gVRF-g association (40.18%) (**Supplementary Table 15**). Second, since myocardial intensity and myocardial volume are associated (**Supplementary Table 11**), it is possible that myocardial intensity is just a measure of myocardial size not well adjusted by regressing out BSA. However, we show that myocardial intensity associates with BMI independent of body and myocardial size (**Supplementary Table 15**). Therefore, myocardial intensity’s mediation of the gVRF-g association is not just explained by the BMI - g association and, furthermore, the BMI - myocardial intensity association is not just due to the myocardium being larger.

We performed serial mediation modelling of the gVRF-g association, testing each imaging latent as a potential mediator. (A) Schematic for the CMR radiomics modelling procedure. gVRF and g were maintained as the known association, and we iterated over all CMR imaging latent factors. Equations demonstrate the derivation of the direct and indirect effect. (B) Schematic for the brain MRI modelling procedure. (C) Example computation of the measured effects. Confidence intervals reported in **Supplementary Table 14**. (C) The estimates for the direct and indirect effects for all potential mediators, sorted by indirect effect size, closed circles are significant (p<0.05) and open are not. Error bars derived from bootstrapping (see **Supplementary Methods**).

### Latent Multiple Mediation Modelling

The structural functional model argues that heart structural variation impacts cognitive function via its impact on brain structure. To model this within our data, we constructed two related multiple mediation models (**Figure 4**). In the first model, we performed ‘parallel’ multiple mediation that does not account for the heart-brain association. In the second model, we performed ‘sequential’ multiple mediation that does account for the heart-brain association.

**Figure 4:**
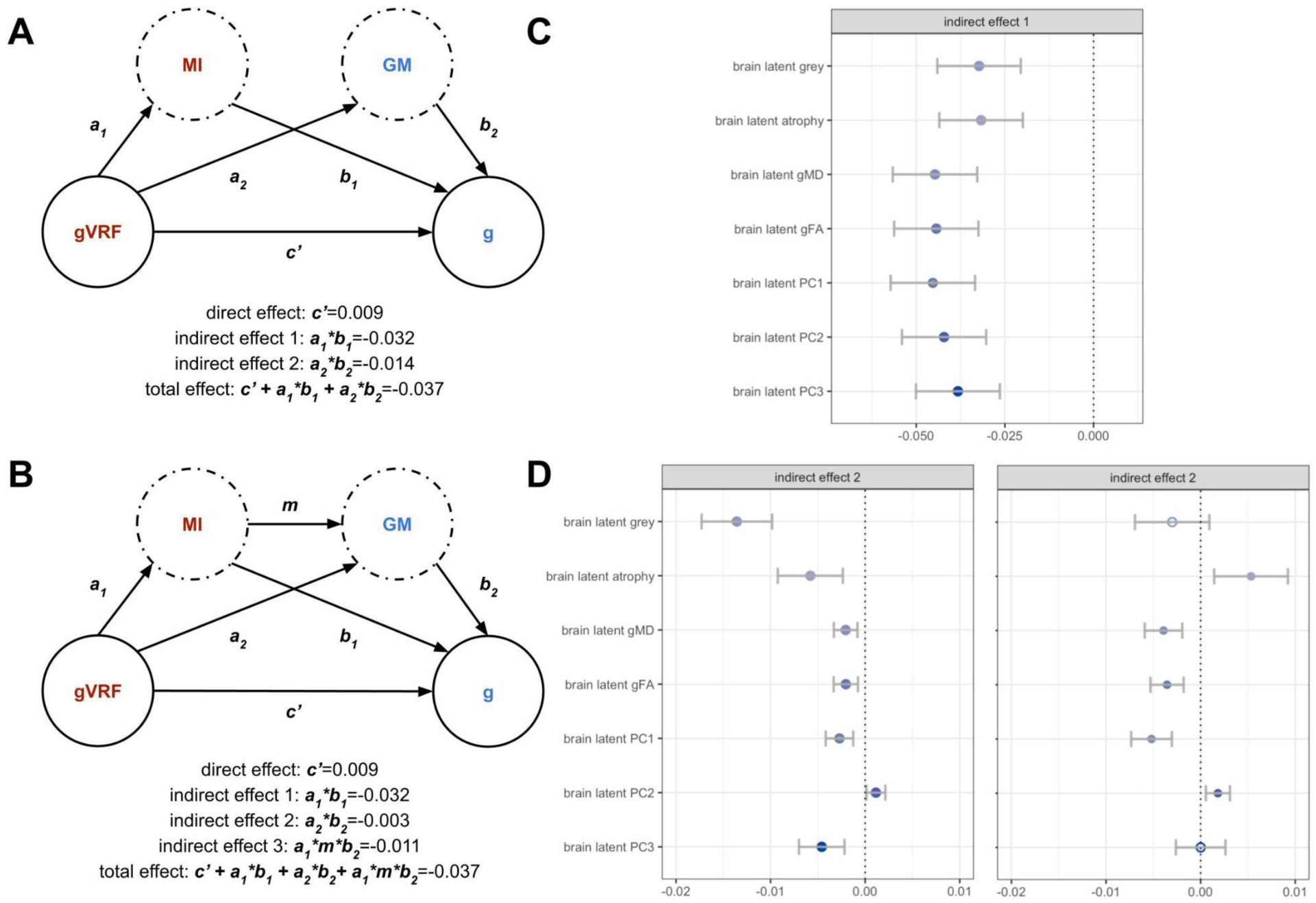
Latent Multiple Mediation Modelling

We performed both parallel and sequential multiple mediation modelling of the gVRF-g association, including heart PC2 as the first mediator and then considering all brain latents as second mediators. (A) Schematic of the parallel modelling procedure with a single direct effect and two indirect effects, one for each potential mediator. We list values from an example mediation effect in which grey matter volume was the second mediator. Confidence intervals reported in **Supplementary Table 17**. The direct effect is fixed for all mediators at 0.009. (B) Analogous schematic for the sequential modelling procedure. Values reported from an example model with grey matter as the second mediator. (C) A bar chart of the estimates for the indirect effect for heart PC2 when each brain latent was used, closed is significant (p<0.05) and open is not. (D) A bar chart of the estimates for the indirect effect for each brain latent when either using parallel (left) or sequential (right) mediation.

We performed this analysis for myocardial intensity paired with every brain latent since myocardial intensity was the only significant heart mediator. Comparing the heart indirect effects between the single mediation (**Figure 3**) and the parallel multiple mediation (**Figure 4C, Supplementary Tables 16, 17**) allows us to assess the impact of brain structure on the heart-cognitive function association. The heart indirect effect slightly decreases when accounting for brain volumes but not brain white matter measures. Therefore, brain volume variation can explain some but not all of the association between heart intensity and cognitive function.

Comparing the brain indirect effects between the parallel and sequential mediation allows assessment of the impact of heart intensity on the VRF-brain association. In this case, the brain volume measure indirect effects go to zero but the white matter indirect effects do not decrease (**Figure 4D, Supplementary Tables 18, 19**). Thus, heart intensity variation can explain all of the association between VRFs and brain volume but not VRFs and white matter intensity.

### Latent Single Mediation Modelling of Individual VRF-Cognitive Pairs

Since recent work has noted the potential for spurious mediations when modelling with composite measures, we spend the next two sections analysing mediation using combinations of individual measures. We first consider pairs of individual VRFs and cognitive exams (**Figure 5, Supplementary Tables 20, 21**). We found that pack years and VNR (β =-0.028), WHR and VNR (β =-0.061), and WHR and RT (β =0.032) all had independent associations in the expected directions. Brain volumetric latents most strongly mediated the pack year - VNR association (12.11–47.64%) while myocardial intensity associated latents most strongly mediated the WHR-VNR association (27.33–42.76%). The myocardial intensity features are also the only significant mediators of the WHR-RT association (34.75–49.52%). Likely because they capture some relevant variation in brain volumes, white matter tracts, and myocardial intensity, the components of the second joint factor strongly mediate both the pack-year and WHR cognitive exam associations (21.63–49.52%).

**Figure 5:**
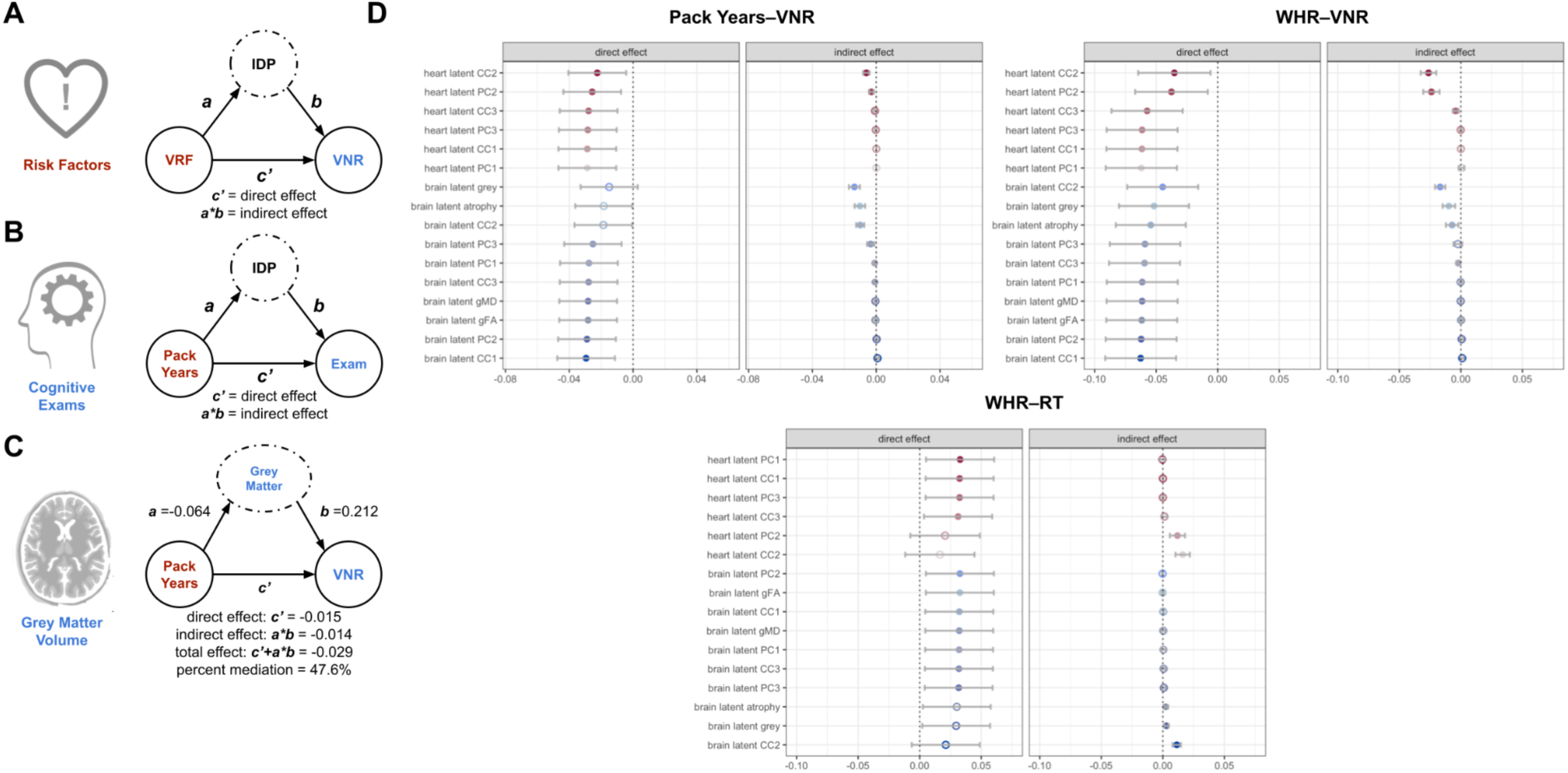
Latent Single Mediation Modelling of VRF-Cognitive Pairs

We performed serial mediation modelling for all latent imaging measures for each VRF-cognitive exam pair. (A) Schematic for a mediation model using different VRFs as the independent variable rather than gVRF. We tested all VRFs. (B) Schematic for a mediation model using cognitive exams as the dependent variable rather than g. We tested all exams. (C) Example mediation model for an individual latent factor and an example pair of VRF and cognitive exam. Confidence intervals for all coefficient estimates in **Supplementary Table 21**. (D) Direct and indirect effects for three significant VRF-exam pairs. Latents ordered by indirect effect size and separated by organ. RT shows lower values for better performance while VNR shows higher values for higher performance.

### Individual Feature Mediation Modelling

Although the latent imaging features capture large amounts of the variance in the imaging datasets (**Supplementary Figure 4, Supplementary Figure 6**), each imaging dataset contains many features and much variance beyond the latents used in the previous analyses. To offer a comprehensive picture of how heart and brain structure mediate the gVRF-g association, we perform single mediation analysis for every individual imaging feature (**Figure 6, Supplementary Tables 22, 23, 24, 25**).

**Figure 6:**
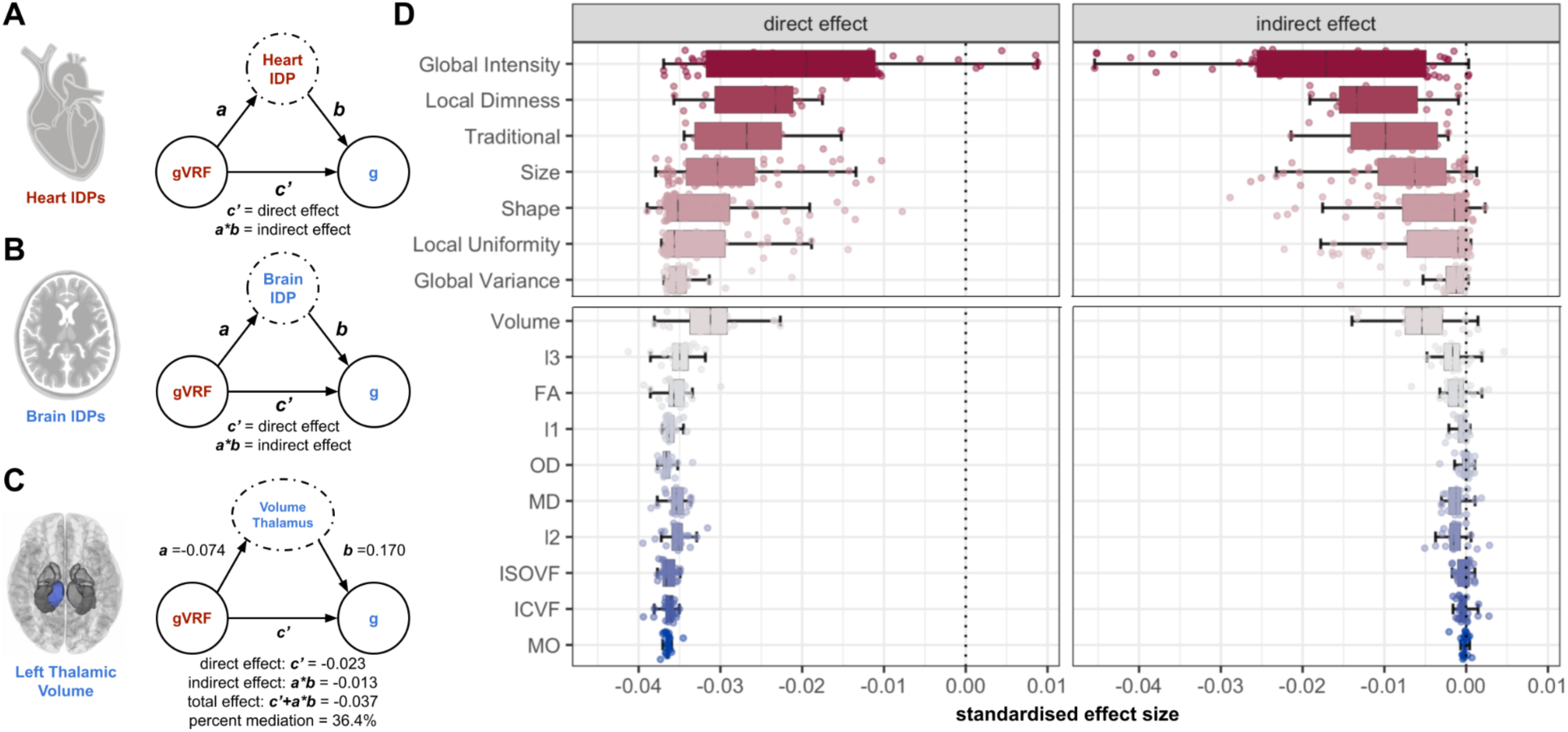
Individual Feature Single Mediation Modelling

As expected, many individual features associated with myocardial intensity show complete mediation (**Figure 6, Supplementary Tables 22, 23**). However, a number of CMR measures showed mediating effects that were previously difficult to appreciate via latent modelling. While the latent measure of myocardial volume did not mediate the association (**Figure 3**), both the right and left ventricular volumes partially mediated the association (32.5–61.1%). Although the latent measure of myocardial tissue complexity was just below significance (**Supplementary Table 14)**, some measures of local nonuniformity and local homogeneity partially mediated the association (32.5–48.5%). Greater local nonuniformity associated with lower vascular risk (β = -0.347– -0.284) and greater cognitive function (β = 0.051–0.054), and measures of local homogeneity show the opposite associations (**Supplementary Table 23**).

Compared to the heart, the brain IDPs show an order of magnitude lower indirect effects and proportionally lower percent mediation (**Figure 6, Supplementary Tables 24, 25**). Of the brain IDPs, volumes have the largest mediating effect, particularly grey matter (38.1%) and thalamic volume (35.9–36.4%). The largest white matter microstructural mediating effects are from the thalamic radiation tracts (**Supplementary Figure 9**). For example, MD of all the thalamic radiation tracts significantly mediates the association (5.17–8.26%), and the FA of the left posterior thalamic radiation tract has the greatest mediation of all the white matter microstructural mediating effects (18.3%).

We performed serial mediation modelling for all individual imaging features. (A) Schematic for the modelling procedure for CMR IDPs. Same as Figure 3, except all potential mediators are now individual features (IDPs) instead of latent variables. (B) Schematic for Brain IDPs. (C) Example mediation model for an individual feature with the left thalamic volume (Volume Thalamus) as a potential mediator. Confidence intervals reported in **Supplementary Table 25**. (D) Direct and indirect effects for all tested CMR radiomics grouped by cluster and brain MRI IDPs grouped by their feature type. For visualisation, we grouped the brain IDPs by their IDP categories and the CMR radiomics by previously reported clusters extracted from imaging of healthy individuals.^18^ We also include conventional CMR indices as a separate cluster.

## Discussion

### Interpretation

This study supports the structural-functional model for the link between vascular risk and cognitive function. Many heart and brain structural measures separately mediate the vascular risk -cognitive function association (**Figure 3, 6**). Despite these initial results, it was still possible that the mediating measures from the heart and brain shared no inter-associations. This would violate the structural-functional model’s claim that vascular risk causes vascular and cardiac remodelling, which in turn causes cerebral damage. Our results argue against this possible negative result in three ways. First, we find significant associations among the separate heart and brain mediators, like between heart PC2 and grey matter volume (**Figure 2**). Second, we find that these associated mediators have partially overlapping mediating effects (**Figure 4**). Third, we find that one of the major axes of covariance between heart and brain structure (CC2) significantly mediated the VRF-CF association (**Figure 3**). Therefore, the heart and brain do indeed share mediating effects, indicating that their variation may be linked via the structural-functional model.

Our results also complicate the structural functional model. When comparing the multiple mediation models (**Figure 4**), we found that myocardial intensity variation can fully explain the VRF-grey matter association, but grey matter variation cannot fully explain the myocardial intensity - cognitive function association. This suggests that heart structural variation can associate with cognitive function in ways independent of brain structural variation. This violation of the structural functional model could be explained by brain changes not well captured by our metrics (e.g. smaller cortical grey matter changes). Additionally, we found that myocardial intensity cannot explain the VRF-white matter integrity association (**Figure 4**). Therefore, the brain associates with risk factors in manners independent of cardiac variation. Mechanisms for this break in the model could be explained by direct impact of metabolic hormonal dysregulation on the brain or brain vasculature, without affecting heart structure.

Consistent with prior reports, considering brain structural measures alone only accounted for a minority of the VRF-CF association.^17^ Although important features from these brain latents all have relatively large coefficients in the second heart-brain CCA mode (**Supplementary Table 7**), these latents alone show much smaller indirect effects than the second heart-brain CCA mode (**Supplementary Table 14**). The strong alignment of the second CCA mode with the VRF-CF association suggests that leveraging the association between heart and brain structure is informative to deriving a brain imaging latent factor that associates with both vascular risk and cognitive decline. In other words, without considering vascular risk or cognitive function in their derivation, one can discover brain biomarkers that better explain the VRF-CF association by using the heart-brain structural association.

Focusing on the brain structures identified, this work unifies separate findings that have shown that lower grey matter and thalamic volume associates with greater vascular risk and lower cognitive function.^17,19,22,26^ Furthermore, this work supports the association of deteriorating thalamic tract white matter microstructure with elevated vascular risk and poorer cognitive function.^19,26^ Previous work has argued that the thalamus is both central to integrative signalling in the brain and potentially susceptible to changes in cerebrovascular perfusion.^29–32^ Crucially, this works links variation in these structures to myocardial intensity. Why exactly thalamic volume and thalamic white matter integrity associate with myocardial intensity is still unknown and will be of interest in future work.

Our analyses of individual VRFs and cognitive exams revealed subtle trends not apparent in our more global/latent results, where brain and heart had differential importance. For example, whereas brain volumes more strongly mediate the pack year - VNR association than the WHR-VNR association, myocardial intensity exhibited the reverse pattern (**Figure 5, Supplementary Table 21**). This result highlights the utility of a comparative approach between heart and brain structural variation. However, the individual VRF cognitive exam analysis also revealed the complexity in some of these phenotypes, replicating a previous finding of a positive association between BMI and visual memory (**Supplementary Table 21**).^17^

Beyond supporting findings from the latent analysis, the individual gVRF-g mediation analysis of imaging features revealed that lower right and left ventricular volume for body size associates with greater vascular risk and lower cognitive function (**Figure 6, Supplementary Table 23**).

This result could point to a simple mechanistic step in the structural-functional hypothesis in which lower stroke volume for body size decreases cerebral perfusion.^28^ Analysis of the individual brain features highlights grey matter, some subcortical volumes, and thalamic white matter tract measures as most mediating the gVRF-g association (**Figure 6, Supplementary Table 25**). This provides independent support from the joint analysis that these specific brain structures are key to the heart-brain axis.

Lower myocardial intensity has previously been associated with specific vascular risk factors and greater red meat consumption, and, here, we quantify its association with both greater aggregate vascular risk and lower cognitive function.^18,33,34^ Lower myocardial intensity strongly associates with higher BMI in a manner not explained by body or heart size (**Supplementary Table 15**), possibly representing fatty transformation of myocardial tissue. Alternatively, previous imaging studies have detected myocardial fibrosis in cohorts of patients with vascular risk factors, suggesting that the low intensity features common to vascular risk and cognitive decline could be signs of a common myocardial fibrotic pathology driven by vascular risk factors.^35–37^ We also found some mediation via greater myocardial textural uniformity (**Supplementary Table 23**), which could also associate with the speculated fibrosis. These results motivate further work to confirm these hypotheses through tissue pathology.

### Limitations

Although this study uses an exceptionally large dataset of adults across a wide range of middle- and older-ages, this work does not analyse longitudinal data. Therefore, we cannot disambiguate whether cardiovascular risk is causing decreased cognitive function, lower cognitive function is causing increased cardiovascular risk, or some mix of both effects. However, numerous longitudinal studies in other cohorts support that cardiovascular risk associates with accelerated cognitive decline.^9–12,38,39^ Furthermore, without longitudinal imaging, we cannot assess the temporal relationship between cardiac and brain imaging phenotypes, vascular risk, and cognitive function. However, we argue that our results still offer novel cross-sectional support for the structural-functional model linking elevated vascular risk and poorer cognitive function.

In this work, we focus on the structural functional model linking vascular risk and cognitive function. Importantly, the VRF - CF association could be equally well explained by unmeasured mechanisms (e.g. metabolic hormonal dysregulation could directly impact neuronal function)^40^ or by reverse causation (e.g. poor cognitive function could decrease healthy lifestyle maintenance).^41,42^ Testing these hypotheses adequately would require longitudinal and biochemical data not yet available via the UK Biobank. Additionally, we do not adjust for ethnicity in this study due to the low numbers of non-White British participants and the heterogeneity of those minority participants (**Supplementary Table 1**).

Whereas some have questioned the reliability of the UK Biobank cognitive exams,^27^ recent work has supported their validity and psychometric properties.^43^ Additionally, as reported in previous work, the effect sizes for the association between individual VRFs and cognitive exams is small, and we find no unique association for many VRFs and at least two associations pointing in the ‘opposite direction’ as hypothesised (**Figure 5, Supplementary Tables 21**).^7^ Results from the full UK Biobank study suggest that large studies are needed to consistently detect these small effects and future increases to the imaging subset will help refine our results.^17,24^ We argue that the approach implemented here, via obtaining a latent measure g, minimises the impact of individual exam variability by obtaining an estimate of a robust, replicable, and test-invariant cognitive construct.^26,27,43^

## Conclusion

The structural-functional model explaining the VRF-CF association rests on the argument that vascular risk drives changes in cardiovascular structure that lead to alterations in brain structure that lead to cognitive decline. Definitive support for the causal sequence of this model would require experimental or longitudinal work. However, our models (using cross-sectional data) are consistent with the hypothesis that vascular risk-associated cognitive ageing associates with distinctive variation in cardiac and brain structure. This is the first large-scale work to show that there is correlated variance in both heart and brain structure that mediates the association between vascular risk and cognitive function, providing a more extensive multi-modal framework to important prior work.^7,17–22,24,26^ One of the many hypotheses generated from analysing these data together is the identification of a key link to explain: how myocardial hypointensity could associate with cerebrovascular hypoperfusion impacting particular subcortical structures, like the thalamus.

## Methods

### Acquisition and Processing

#### Assessment

This work utilises clinical and imaging data from the United Kingdom (UK) Biobank via access application 2964.^44^ The UK Biobank is a large-scale longitudinal dataset derived from 500,000 volunteers recruited between 2006 and 2010 from across the UK. At visits, participants completed both a touchscreen questionnaire and medical history interview with a nurse. The project recorded information regarding participants’ health, lifestyle, and family history and collected physical measurements, biological samples, and genome. Moreover, since 2015, over 50,000 participants have received CMR and brain MR imaging at followup imaging visits.

#### Vascular Risk Factors

We analysed hypercholesterolemia, diabetes, hypertension, smoking pack years, blood pressure, and anthropomorphic measures (BMI and waist-to-hip ratio, WHR).^3,13–15,19,45^ All vascular risk factors were collected at the baseline UK and prepared as reported previously.^18,19^ We summarise the process here. During the medical history interview, participants reported whether they had received a diagnosis of diabetes, hypertension, or hypercholesterolaemia. Diagnosis was confirmed through a combination of HES records and blood biochemistry data.^18^ Participants provided information on cigarette smoking in the touchscreen questionnaire, and smoking pack years were computed from this data.^19^ Blood pressure was collected twice, moments apart, using an Omron 705IT monitor. Mean systolic and diastolic blood pressure were computed.

Anthropometric measures were taken after participants had removed bulky clothing and shoes. Waist and hip measurements were conducted to provide WHR. BMI was computed by dividing weight by squared height.

#### Cognitive Exams

We examined four tests that were included as part of the UK Biobank baseline cognitive assessment. The complete battery and assessment of its repeatability and reliability have been detailed previously.^7,27,43^ We used the four tests commonly used in analysis and dimensionality reduction of the baseline cognitive assessment: the fluid intelligence task (verbal numerical reasoning, VNR), the visual memory task (vismem), the reaction time task (RT), and the prospective memory task (prosmem).^27^ As previously reported,^27^ the reaction time scores were positively skewed, so we applied a natural log transformation (LN). Additionally, the visual memory scores were zero-inflated and positively skewed, so we applied a LN+1 transformation.

#### Cardiac Imaging

Cardiac imaging acquisition and preparation discussed in **Supplementary Methods**. Using the CMR images and their corresponding segmentations, we performed radiomics phenotyping based on the open-source python-based pyradiomics library.^46^ Radiomics extracts features quantifying myocardial and ventricular structure (shape radiomics), myocardial imaging intensity (first-order radiomics), and myocardial visual textures (texture radiomics).^47^ In total, 212 features per region were extracted at end-diastole and end-systole. Right and left ventricular cavity first-order and texture features were excluded from analysis because they do not encompass clinically relevant information. We incorporate conventional CMR indices into the matching analysis and final mediation by individual features, computed as previously reported.^20,21,34^

#### Brain Imaging

Brain imaging acquisition and preparation is discussed in **Supplementary Methods**. The global tissue volumes and white matter tract-averaged water molecular diffusion indices were processed by the UK Biobank team and made available to approved researchers as imaging-derived phenotypes (IDPs); the full details of the image processing and QC pipeline are available in an open access article.^48^ The IDPs in this study included total brain volume, grey matter volume, subcortical volumes, and tract-averaged white matter microstructural measures. A detailed list of volumes, white matter tracts, and white matter tract measures is provided in **Supplementary Methods**.

### Analysis

#### Workflow

We began with 19408 subjects with completed CMR radiomics analysis of their short-axis imaging from the UK Biobank Imaging Extension. We downloaded and prepared the vascular risk factor, cognitive testing, brain imaging data, heart imaging, and covariates for these subjects (see Acquisition and Preparation). For each dataset separately, we dropped all subjects without complete data, merged all datasets, and selected only subjects without cardiovascular or brain disease (defined in **Supplementary Methods**). We then performed dimensionality reduction on each data type separately. We performed joint factorization of the heart and brain imaging data. We regressed out imaging confounders from the latent factors (**Supplementary Methods**). We merged the latent factors and performed all downstream analyses. We corrected all comparisons for multiple hypothesis testing with a Benjamini-Hochberg False Discovery Rate (BH-FDR) correction. Entire pipeline with number of subjects retained at each step reported in **Supplementary Figure 1** and population statistics reported in **Supplementary Table 1**. For every analysis, we present both raw and deconfounded results as paired **Supplementary Tables**, but we only discuss deconfounded results in the text. All code was open-sourced, see **Data and Code Availability**; the list of packages and settings used is in **Supplementary Methods**.

#### Dimensionality Reduction

##### Latent Variables for Vascular Risk (gVRF)

First, we derived an aggregate measure of vascular risk for each individual, counting instances of a self-reported diagnosis of hypertension, diabetes, or hypercholesterolaemia, having ever smoked, having a BMI >25, and having a high WHR (>0.85 for females and >0.90 for males).^19,49^

We derived an additional latent factor of general vascular risk (gVRF) following prior work in this and other cohorts, using confirmatory factor analysis in structural equation modeling.^19,50^ This latent measure captures the tendency for VRFs to co-occur. gVRF was derived from smoking pack years, diastolic and systolic blood pressure, BMI, WHR, self-reported hypertension, diabetes and hypercholesterolaemia. The model fit the data well, though loadings were inconsistent (range 0.189–0.745), with the factor more strongly loaded towards BMI and WHR (**Supplementary Figure 2, Supplementary Table 2**).

##### Latent Variables for Cognitive Function (general intelligence, g)

As previously reported,^7^ we performed a CFA of the four cognitive tests. We hypothesised that the four tests would correlate moderately-highly (with intercorrelations of r > 0.40) and would form a single latent general factor across the four tests with good fit to the data. We found this to be the case (**Supplementary Figure 3, Supplementary Table 3**).

##### Latent Variables for Heart Structure

Since principal component analysis (PCA) is commonly used in radiomics to extract lower dimensional representations of the data,^47,51–53^ we performed PCA on the z-scored radiomics. We chose the number of principal components using cross validation, detailed in the **Supplementary Methods**. We kept the first 3 unrotated PCs (**Supplementary Figure 4, Supplementary Table 4**). We extracted the scores of these components for each subject and used them for downstream analyses.

##### Latent Variables for Brain Structure

We isolated brain volume (‘atrophy’ after controlling for head size) and grey matter volume.^19^ Latent measures of general white matter fractional anisotropy (gFA) and mean diffusivity (gMD) were derived using confirmatory factor analysis, as previously reported in this cohort.^19,32^ The factor analysis models fit well with the lowest loadings for the corticospinal tracts and cingulate gyri and the highest loadings for the thalamic radiata and fasciculi (**Supplementary Figure 5, Supplementary Table 5**).

Since principal component analysis has been used to capture variation in brain imaging in previous work and since we are using it to summarise the heart imaging in this work,^54–57^ we also computed PCA over all z-scored brain IDPs and selected the number of principal components to retain as before (**Supplementary Methods**). We kept three PCs (**Supplementary Figure 6, Supplementary Table 6**). We extracted their scores for each subject and utilised them in downstream analyses.

##### Joint Heart-Brain Factor Analysis

Along with the factor analysis of the individual datasets described above, we also sought to derive latent factors that captured the main modes of correlated variation between heart and brain structural imaging. That is, we aimed to identify components of brain structure and components of heart structure that were maximally correlated. Through canonical correlation analysis (CCA) on the z-scored heart radiomics and brain IDPs, we derived ten modes.^55^ Each mode consists of two components: (1) a linear combination of heart radiomics features and (2) a separate linear combination of brain IDPs that have highly similar variation in the population. The modes are ranked by the amount of correlation between the heart and brain components. We chose the number of modes to keep via cross validation (**Supplementary Methods**), kept three modes (**Supplementary Figure 7, Supplementary Table 7**), extracted the component scores for each subject in each dataset, and used them in downstream analyses.

#### Descriptive Statistics and Associations

First, we conducted descriptive analyses, testing the association of age and sex with all of our latent variables using linear regression. We then examined the pairwise linear association between all latent variables by linearly modeling each latent variable as a function of sex, age, and each other latent variable. See **Supplementary Methods** for modelling details and how additional R^2 is computed. Results reported for both raw and deconfounding imaging latents.

#### Propensity Score Matching

Since all other analyses are performed on corrected, standardised, and latent measures of the data, we performed propensity score matching to yield real-units measurements of the differences between subjects with and without VRFs. We matched subjects with four or more VRFs with their nearest neighbour with no VRFs, requiring an exact match for sex (**Supplementary Methods**). We then performed repeated t-tests to compare the cognitive exam performance, CMR measures, and brain IDPs of the matched groups of subjects.

#### Mediation Modelling

To measure how well heart and brain structural features explain the VRF-CF association, we perform a series of mediation analyses. This method allows us to directly quantify the degree to which any identified associations between vascular risk and cognitive ability are accounted for by brain or heart-based measures. The primary outcome is therefore the % of the gVRF-g association that is mediated when brain/heart measures are included in the model. In more complex models with more than one mediator, one can also identify which mediator is contributing the largest unique mediating effect. Thus, these analyses offer an elegant quantitative solution for identification of important heart and brain biomarkers underpinning VRF-cognitive associations. We report a more complete description of the mediation model in the **Supplementary Methods**.

We first performed mediation models on solely the latent representations of each data set. We found the association between gVRF and g and then modelled how well each imaging latent variable mediated this association (more details in **Supplementary Methods**). At first, we only modelled one imaging latent at a time, calling this the ‘Latent Single Mediation Model.’ Then we performed both parallel and sequential multiple mediation analyses, fixing heart PC2 as the first mediator and then adding brain latents as the second mediator, called ‘Latent Multiple Mediation Model.’ Next, we replaced the gVRF-g association with pairs of individual VRFs and cognitive exams, testing imaging latents one at a time again, calling this ‘Latent Single Mediation Modelling of VRF-Cognitive Pairs.’ Given the high association between the VRFs (**Supplementary Figure 2, Supplementary Table 2**), we control each VRF-exam association for all other VRFs to identify unique associations between each VRF and cognitive exam.

To explore the role of individual imaging features in explaining the association between VRFs and CF, we returned to the gVRF-g association and performed mediation modelling for each imaging feature individually, calling this the ‘Individual Feature Single Mediation Model.’ We perform modelling as described in **Supplementary Methods**.

Given that all latent measures across domains (vascular risk, heart, brain and cognitive) were standardised, reported coefficients are standardised regression coefficients (i.e. β range [-1, 1]) throughout, allowing direct comparison of effect magnitudes across modalities.

## Data Availability

All data used in the present study are available upon application from the UK Biobank

https://www.ukbiobank.ac.uk/

## Acknowledgments

We thank the UK Biobank participants and the UK Biobank team for their work in collecting, processing and disseminating these data for analysis. AJ received funding from a Fulbright Pre-doctoral Research Award (2019–2020). This research was funded in whole, or in part, by the Wellcome Trust [221890/Z/20/Z and 108890/Z/15/Z]. For the purpose of open access, the author has applied a CC BY public copyright licence to any Author Accepted Manuscript version arising from this submission. ELSC is supported by funding from the Wellcome Trust 4-year PhD in Translational Neuroscience (108890/Z/15/Z).ZR-E recognises the National Institute for Health Research (NIHR) Integrated Academic Training programme which supports her Academic Clinical Lectureship post and was also supported by British Heart Foundation Clinical Research Training Fellowship No. FS/17/81/33318. Barts Charity (G-002346) contributed to fees required to access UK Biobank data [access application #2964]. SEP acknowledges the British Heart Foundation for funding the manual analysis to create a cardiovascular magnetic resonance imaging reference standard for the UK Biobank imaging resource in 5000 CMR scans (www.bhf.org.uk; PG/14/89/31194). SEP acknowledges support from the National Institute for Health Research (NIHR) Biomedical Research Centre at Barts. PG, KL and SEP have received funding from the European Union’s Horizon 2020 research and innovation programme under grant agreement No 825903 (euCanSHare project). SEP acknowledges support from and from the “SmartHeart” EPSRC programme grant (www.nihr.ac.uk; EP/P001009/1). SEP also acknowledges support from the CAP-AI programme, London’s first AI enabling programme focused on stimulating growth in the capital’s AI Sector. CAP-AI is led by Capital Enterprise in partnership with Barts Health NHS Trust and Digital Catapult and is funded by the European Regional Development Fund and Barts Charity. This article is supported by the London Medical Imaging and Artificial Intelligence Centre for Value Based Healthcare (AI4VBH), which is funded from the Data to Early Diagnosis and Precision Medicine strand of the government’s Industrial Strategy Challenge Fund, managed and delivered by Innovate UK on behalf of UK Research and Innovation (UKRI). Views expressed are those of the authors and not necessarily those of the AI4VBH Consortium members, the NHS, Innovate UK, or UKRI. This work was supported by Health Data Research UK, an initiative funded by UK Research and Innovation, Department of Health and Social Care (England) and the devolved administrations, and leading medical research charities. This project was enabled through access to the MRC eMedLab Medical Bioinformatics infrastructure, supported by the Medical Research Council (www.mrc.ac.uk; MR/L016311/1). SRC is supported by a Sir Henry Dale Fellowship, jointly funded by the Wellcome Trust and the Royal Society (221890/Z/20/Z), and acknowledges funding from Biotechnology and Biological Sciences Research Council, and the Economic and Social Research Council (BB/W008793/1), Age UK (The Disconnected Mind project), the US National Institutes of Health (R01AG054628; 1RF1AG073593), the Medical Research Council (MR/R024065/1), and The University of Edinburgh. KL received funding from the Spanish Ministry of Science, Innovation and Universities under grant agreement RTI2018-099898-B-I00.

## Data and Code Availability

UK Biobank Data is available via application. All code open-sourced here: https://github.com/akshay-jaggi/heart_brain_mediation

## Contributions

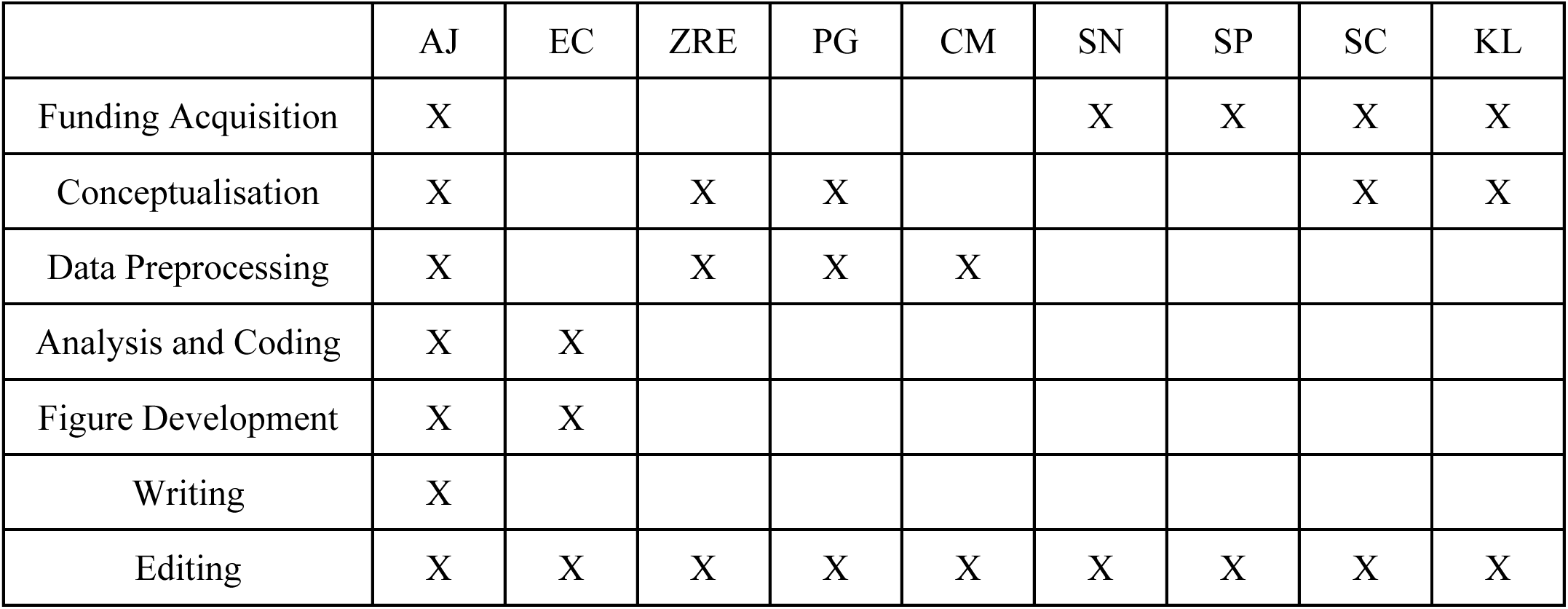

