## Supplementary Methods and Figures for "A Structural Heart-Brain Axis Mediates the Association Between Cardiovascular Risk and Cognitive Function"

#### Acquisition and Processing

##### Heart Imaging

The UK Biobank acquired all images according to predefined standardised operating procedures.^1,2^ All CMR images were acquired using 1.5 Tesla scanners (MAGNETOM Aera, Syngo Platform VD13A, Siemens Healthcare, Erlangen, Germany) following the standardised protocol. We analysed the complete short axis stack covering the left and right ventricles acquired at one slice per breath hold using balanced steady-state free precession sequences. We took images from end-systole and end-diastole during this sequence. These images were segmented through a combination of manual and automated procedures detailed previously.^3–5^

##### Brain Imaging

All brain MRI data were acquired on the same model of 3T Siemens Skyra scanner, according to a freely available protocol.^6^ All imaging parameters have been reported previously.^7,8^ T1-weighted MPRAGE and T2-weighted FLAIR volumes were acquired in sagittal orientation at 1 × 1 × 1 mm and 1.05 × 1 × 1 mm resolution, respectively. The dMRI acquisition comprised a spin-echo echo-planar sequence with 10 T2-weighted (b ≈ 0 s mm^−2^) baseline volumes, 50 b = 1000 s mm^−2^ and 50 b = 2000 s mm^−2^ diffusion weighted volumes, with 100 distinct diffusion-encoding directions and 2 mm isotropic voxels.

The IDPs in this study included total brain volume, grey matter volume, subcortical volumes (accumbens, amygdala, caudate, hippocampus, pallidum, putamen, thalamus), and tract-averaged fractional anisotropy (FA), mean diffusivity (MD), intra-cellular volume fraction (ICVF), isotropic or free water volume fraction (ISOVF), and orientation dispersion index (OD) of the following white matter tracts: acoustic radiation, anterior thalamic, cingulum gyrus, and parahippocampal, corticospinal, forceps major and minor, inferior fronto-occipital, inferior longitudinal, middle cerebellar peduncle, medial lemniscus, posterior thalamic, superior longitudinal, superior thalamic, and uncinate.

#### Analysis

##### Workflow

###### Heart Disease Filtering

To filter out patients with severe cardiovascular pathology, we identified all patients with any ischaemic heart disease, non-ischaemic cardiomyopathy, valvular disease, or significant arrhythmia. We derived these diagnoses through a combination of self-reported answers at baseline interview, UK Biobank algorithmically-computed outcomes, and linked HES data codes, reported in detail previously.^9^ 1311 of the 13709 subjects that passed initial filtering for completeness had at least one of the above conditions.

###### Brain Disease Filtering

To align with previous studies and observe only non-pathological variation in cognitive function and brain structure,^8^ we removed all patients with the following diseases provided via self-report during the baseline UK Biobank session: dementia, parkinsons, other chronic/ neurodegenerative, Guillain-Barré, multiple sclerosis, other demyelinating, stroke, brain haemorrhage, brain/intracranial abscess, cerebral aneurysm, cerebral palsy, encephalitis, epilepsy, head injury, infection of nervous system, ischaemic stroke, meningioma (benign), meningitis, motor neuron disease, neurological injury/trauma, spina bifida, subdural haematoma, subarachnoid haemorrhage, transient ischaemic attack, brain cancer, and meningeal cancer. 524 of the 13709 subjects that passed initial filtering for completeness had at least one of the above conditions.

##### Dimensionality Reduction

###### CMR Radiomics PCA

To select the number of robust principal components to retain for downstream analysis, we ran 10-fold cross validation over the dataset, computed principal components on the training data, computed the explained variance in the held-out data, plotted the explained variance, and found the elbow in this curve, the point after which the explained variance stops decreasing across PCs.

Here we offer interpretations of the loadings of the principal components to understand their significance (**Supplementary Table 4**). The first PC has large weights for myocardial volume, surface area, and several global texture features correlated with myocardial size. The second PC has large weights for measures of the centre of the myocardial voxel intensity distribution (e.g. mean, median). The third PC has large negative weights for measures of the variability of the myocardial textures (e.g. entropy, contrast). Such that the value of the third PC correlates with complexity, we used the negative of its PC scores.

###### Brain MRI IDP PCA Interpretation

For the Factor Analysis, tract measures (left and right) were entered separately into this analysis, correlated residuals between the left and right of each tract and between some other tracts were allowed.

We offer interpretations of the loadings of the first three unrotated principal components of the Brain MRI IDPs (**Supplementary Table 6**). The first PC has high weightings for ICVF and FA of the fasciculi and thalamic tracts. The second PC has large negative weightings for many white matter integrity measures of the corticospinal tracts. We took the negative of the second PC scores such that it increases with white matter integrity. The third has high weightings for volumes, suggesting a factor capturing size.

###### Heart-Brain Joint Factor Interpretation

To choose the number of CCA modes to keep, we performed 10-fold cross validation on the subjects. For each training fold, we computed ten CCA modes and extracted their loadings. We then applied the loadings to the held-out testing data and found the correlation of the heart and brain components in the held-out data. We then plotted the correlation and chose the number of modes to the left of the elbow, or before the correlation in the modes levels off. Like the PCA method, this cross-validation method ensures that we choose a number of features with reasonable explained variance in unseen UK Biobank subjects.

To assess what features are most important to each mode, we found the correlation of the component scores with the features of each dataset (**Supplementary Table 7**). We found that most interesting associations were negative for the first two modes, so we inverted the sign of their component scores for clarity. The scores of the first mode correlate with most volume measures from both the heart and brain imaging. The scores of the second mode correlate with myocardial intensity in end-systole and correlate with some measures of volume of the brain image (notably grey matter volume) and many measures of white matter integrity (ICVF and FA) in the thalamic tracts. We expect both volumes to decrease and FA to decrease in neurovascular pathology, so this mode likely correlates with neurovascular health.^8^ The scores of the third mode correlate with myocardial intensity in end-diastole, and they also correlate with MD.

##### Imaging Deconfounding

Both of the imaging datasets generate features sensitive to the size of patients and their position in the scanner. To remove potential confounding on downstream analyses, we regressed all latent variables for the imaging datasets on the imaging confounders and performed future analyses on the residuals of this regression. For the cardiac imaging latents, we regressed each latent onto z-scored body surface area (BSA). We regressed each brain imaging latent onto z-scored head size and the head position in the scanner (X, Y, and Z coordinates). We regressed the components of the joint factors on both the heart and brain imaging confounders because the joint analysis can induce correlations with imaging conditions from either dataset. In analyses that use individual imaging features, we perform this deconfounding on individual features rather than the latent features. Since the uncorrected values may be of interest in some contexts, we report both the uncorrected and corrected values for all analyses in the **Supplementary Tables**.

##### Linear Models

In **Figure 3, Supplementary Tables 9, 10**, we report the results of linearly modeling each latent measure as a function of each other latent measure controlling for sex and age. We report additional R^2 as the amount of variance that the independent latent measure explains in the dependent latent measure above what is already explained by the covariates.

##### Mediation Modelling

Since mediation modelling has been described in detail elsewhere,^10^ we will provide a brief explanation of the procedure in the context of this study. We first regress a potential mediator, say grey matter, on gVRF. Label this coefficient ***a***. We then regress g on both the mediator and gVRF. The coefficient for the mediator is ***b***, and the coefficient for gVRF is ***c’***. The indirect effect of gVRF on g via the mediator is ***a*b***, and the direct effect of VRF on g is ***c’***. The magnitude of the indirect effect indicates the degree to which a mediator explains the observed association between gVRF and g. The total effect is ***c=c’+a*b***. The percent mediation is the ratio of the indirect effect and the total effect: ***a*b/(c’+a*b)***.

Along with single mediation, we also performed and report parallel and sequential multiple mediation models. We considered two mediators in each multiple mediation model. The first mediator was always Heart PC2 (mediator 1), and we used the brain latents as the second mediator (mediator 2). In parallel multiple mediation, the mediator 1 is not explicitly considered as a predictor for mediator 2 while it is considered in sequential mediation. Therefore, ***c=c’+a_1_*b_1_+a_2_*b_2_*** in parallel mediation, and ***c=c’+a_1_*b_1_+a_2_*b_2_+a_1_*m*b_2_*** in sequential mediation, where ***m*** is the coefficient of the mediator 2 regressed on mediator 1. Comparing the indirect effect from the parallel multiple mediation to the single mediation allows for one to assess how unique the g associations for mediator 1 and mediator 2 are. Similarly, comparing the parallel multiple mediation and the sequential multiple mediation allows one to assess the uniqueness of the gVRF association for mediator 2. In other words, parallel multiple mediation measures how much mediator 1 mediates the association between gVRF and mediator 2.

#### Packages

For gVRF, g, gMD, and gFA extraction, we performed confirmatory factor analysis (CFA) using ‘cfa’ from the lavaan R package.^11^ For PCA of the CMR radiomics and brain IDPs, we used prcomp of the base R stats package. We performed CCA using ‘CCA’ from the scikit-learn cross-decomposition module in Python.^12^ We performed all linear modelling with lavaan in R and report bootstrapped estimates of confidence intervals, method previously reported.^11^ We control for multiple hypothesis testing across all linear models by performing a Benjamini-Hochberg False Discovery Rate (BH-FDR) adjustment for all latent variable p-values.^13^ We performed propensity score matching using ‘matchit.’^14^ We used a logistic regression distance metric and a calliper of 0.05. We matched men and women separately and then recombined. For the mediation models, we performed fitting and bootstrapping via lavaan in R and report the direct effect, indirect effects, total effect, and percent mediation.^11^ In all models, we adjusted for sex and age, report both raw and deconfounded results, and correct for multiple hypothesis testing via a BH-FDR correction.

### Supplementary Figures

#### Supplementary Figure 1

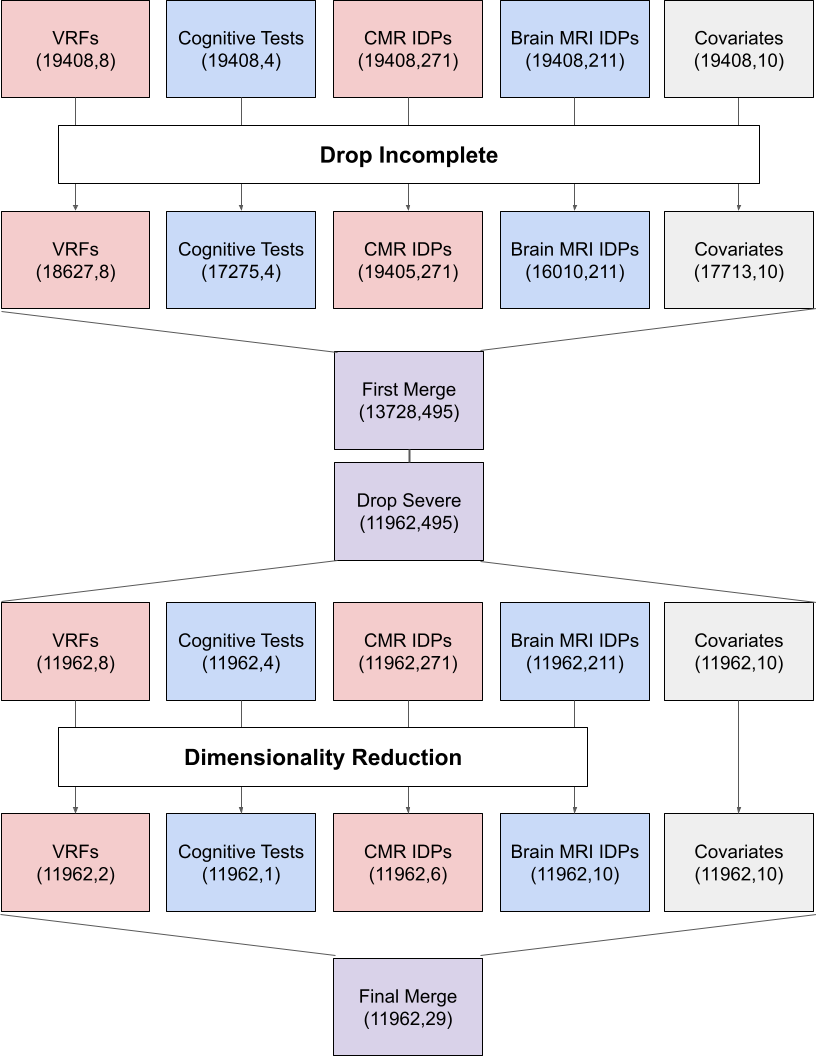

Supplementary Figure 1: Data Preparation Workflow

The flow of subjects and variables through data preparation. Each box represents the number of subjects and the number of variables after the most recent action was taken. We began with 19408 subjects with complete CMR imaging and pulled their data for the other categories. We then dropped incomplete subjects for each category and removed all subjects with a severe cardiovascular or brain disease diagnosis. We then separately conducted factor analysis (and joint factor analysis not illustrated here for clarity) and finally merged all latent variables and covariates for downstream modelling. Characteristics of final cohort in **Supplementary Table 1**.

#### Supplementary Figure 2

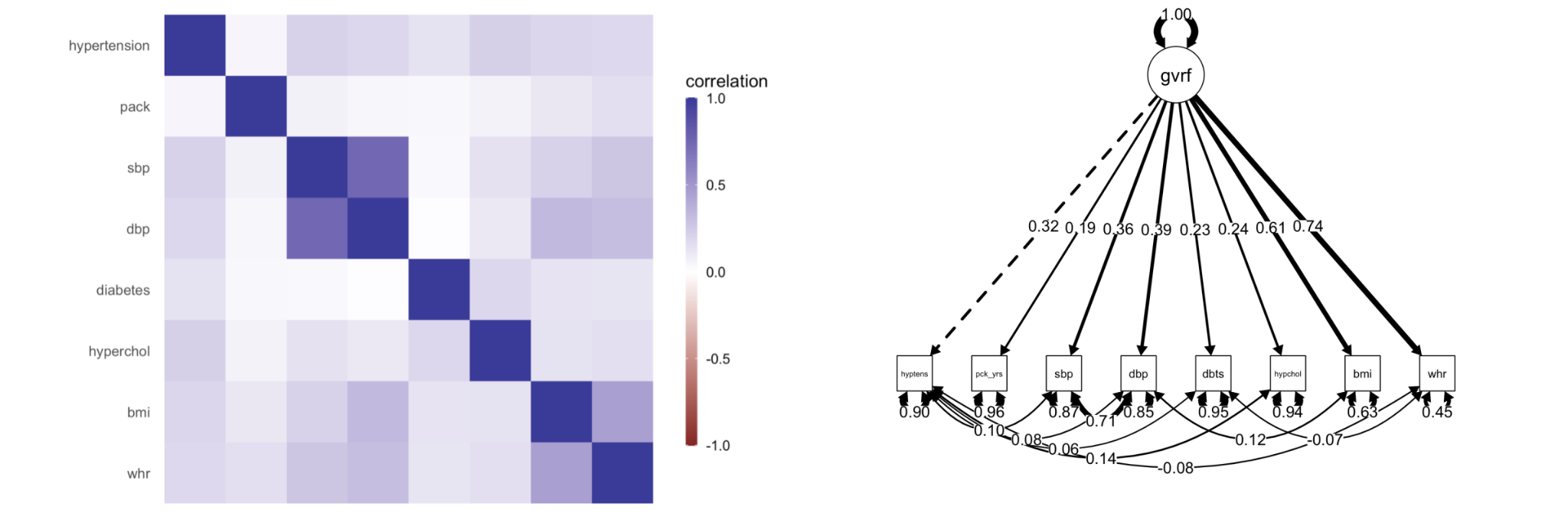

Supplementary Figure 2: CFA for Vascular Risk

A correlation heatmap and factor loadings for all vascular risk variables. Fit measures reported in **Supplementary Table 2**.

#### Supplementary Figure 3

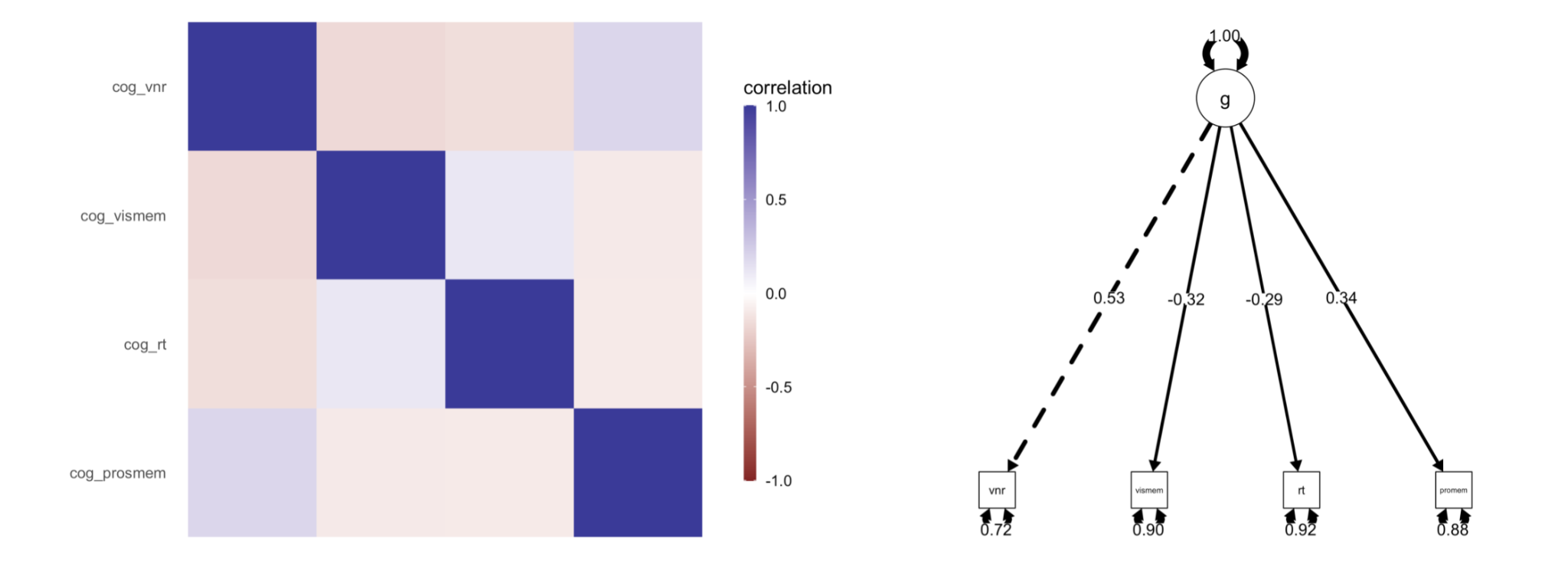

Supplementary Figure 3: CFA for Cognitive Function

A correlation heatmap and factor loadings for the cognitive exams. Fit measures reported in **Supplementary Table 3**.

#### Supplementary Figure 4

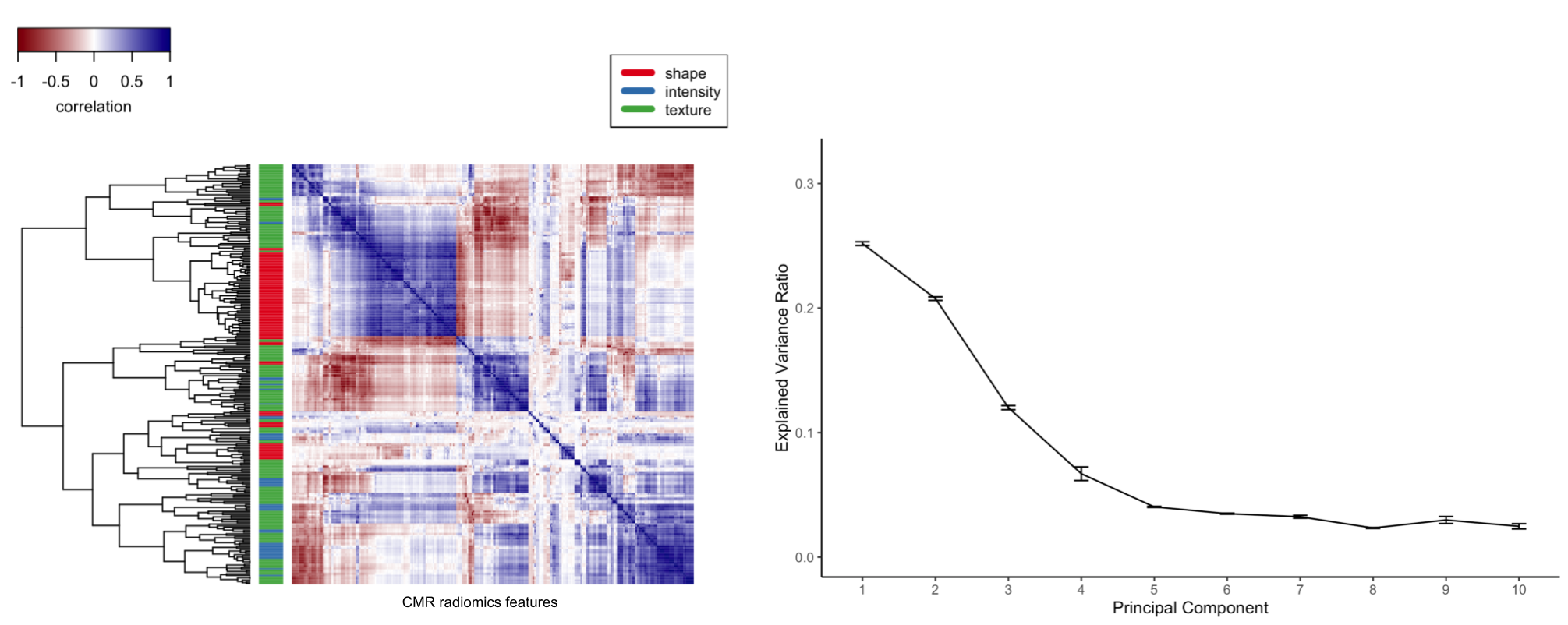

Supplementary Figure 4: PCA for CMR Radiomics

A correlation heatmap of CMR Radiomics features (sorted by a complete linkage dendrogram) with a colorbar for the original feature type. Percent variance explained of the top ten principal components of the features, error bars represent standard error over 10-fold cross validation. Individual feature loadings for each component are reported in **Supplementary Table 4**.

#### Supplementary Figure 5

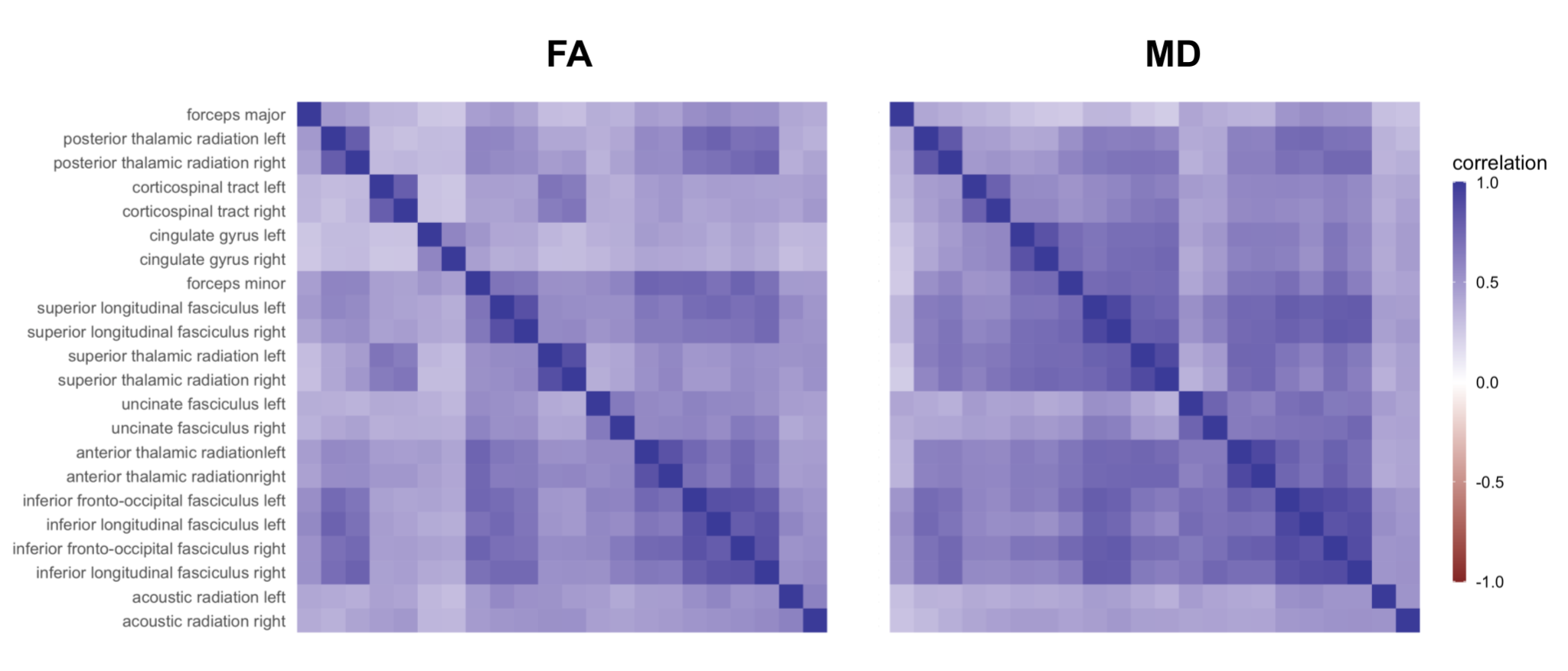

Supplementary Figure 5: CFA for FA and MD

A correlation heatmap of FA and MD IDPs. Fit measures and variable loadings are reported in **Supplementary Table 5**.

#### Supplementary Figure 6

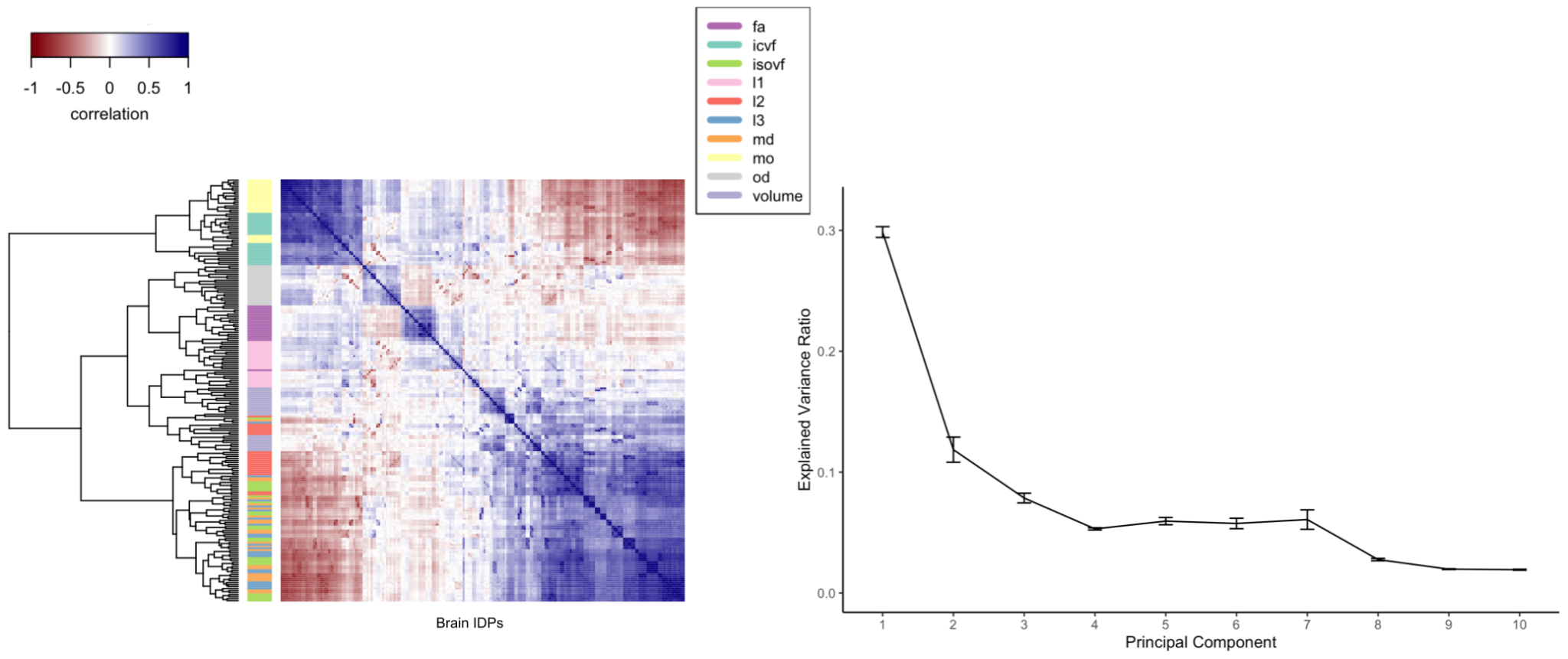

Supplementary Figure 6: PCA for Brain IDPs

A correlation heatmap of brain MR IDPs (sorted by a complete linkage dendrogram) with a colorbar for the original feature type. Percent variance explained of the top ten principal components of the features, error bars represent standard error over 10-fold cross validation. Individual feature loadings for each component are reported in **Supplementary Table 6**.

#### Supplementary Figure 7

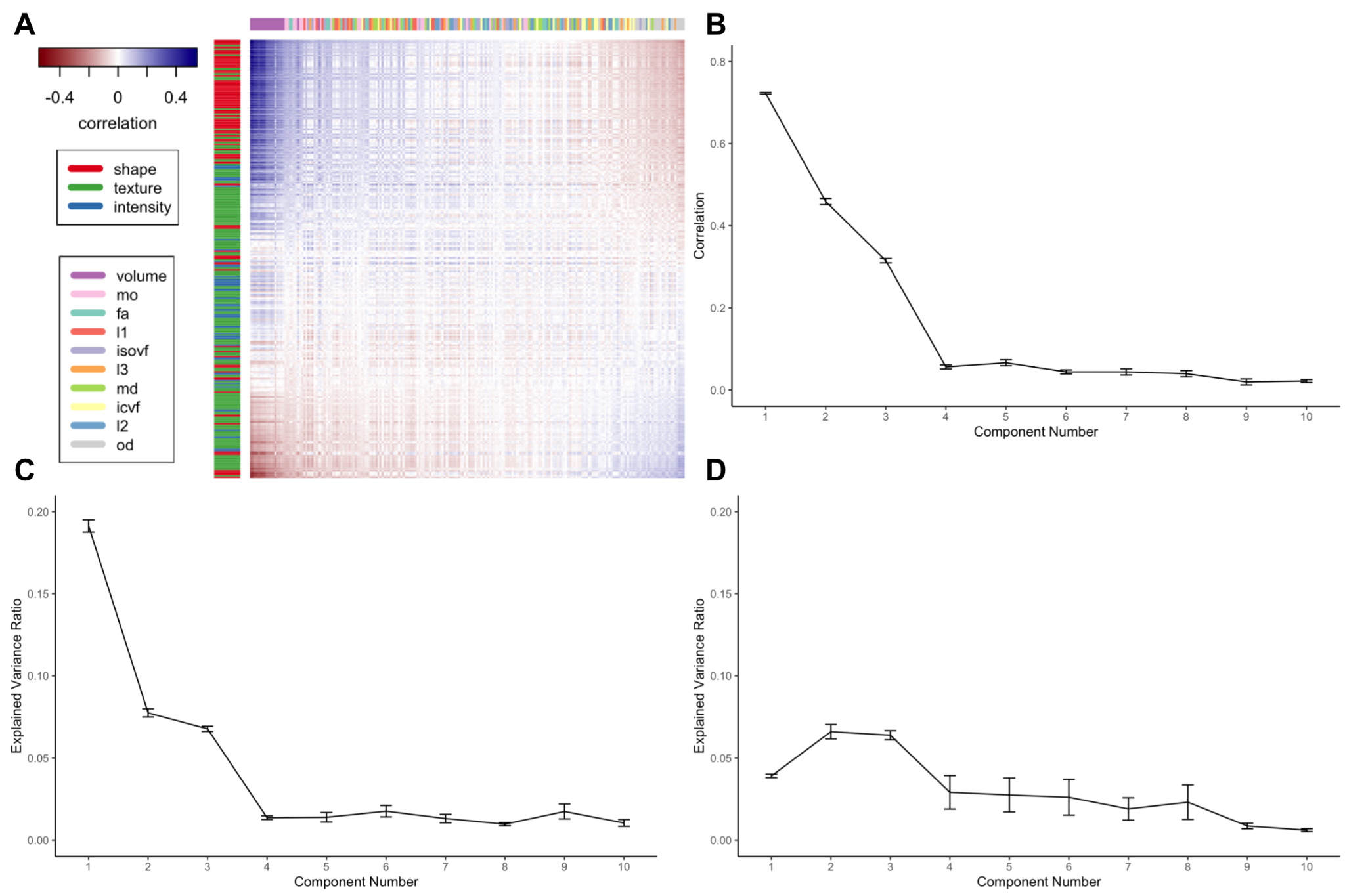

Supplementary Figure 7: CCA for Heart and Brain IDPs

(A) A correlation heatmap of the CMR Radiomics (rows) and Brain IDPs (columns). The heatmap columns and rows have been ordered by the weight of each feature in the loadings of the first CCA mode. (B) The correlation between the first ten CCA modes for the heart and brain imaging data in held-out data. The errors represent standard error over 10-fold cross validation. (C) The variance explained in held-out data by the heart component of each CCA mode. (D) The variance explained in held-out data by the brain component of each CCA mode. Individual feature loadings are reported in **Supplementary Table 7**.

#### Supplementary Figure 8

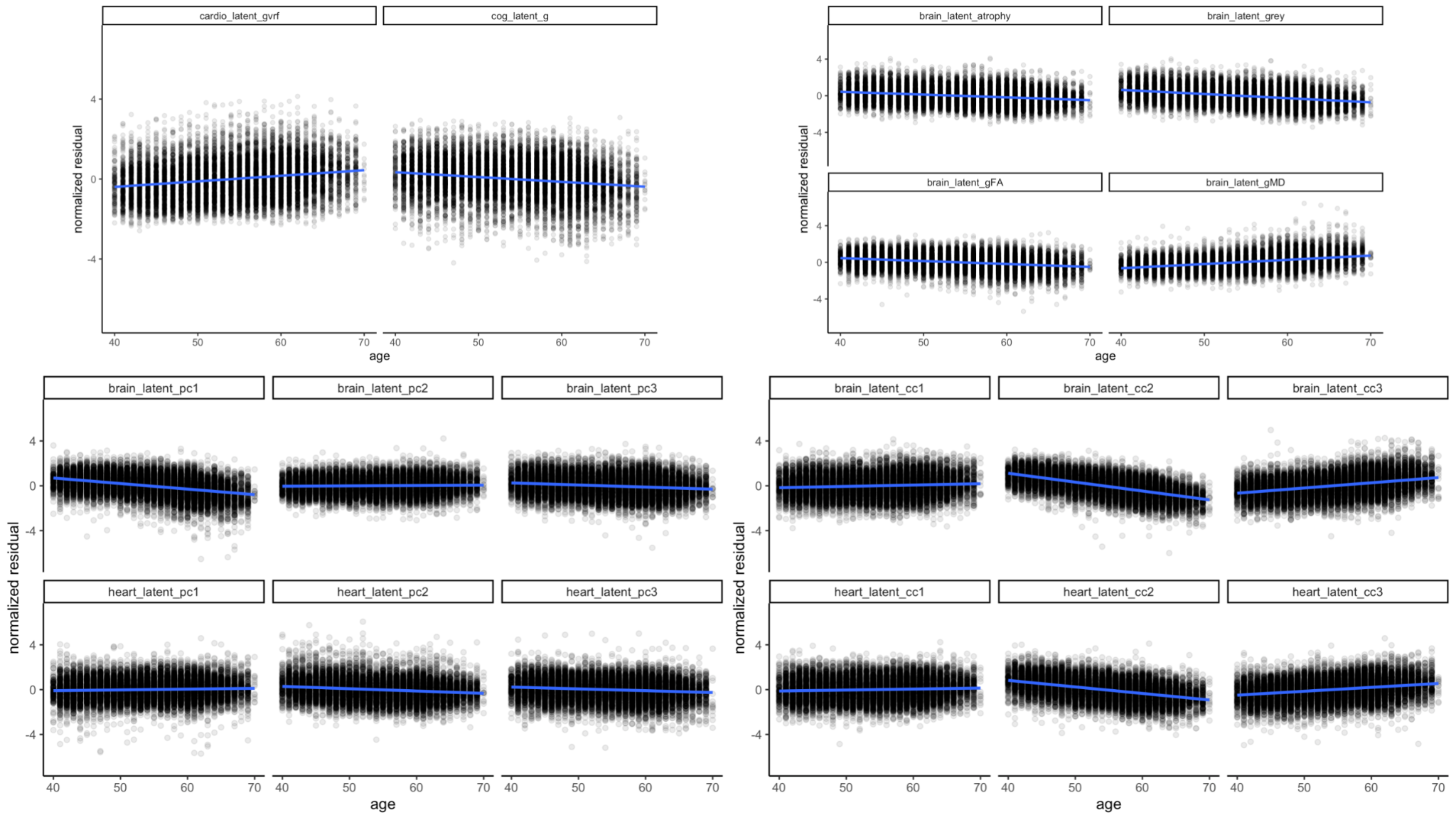

Supplementary Figure 8: Age Associations

Plot of each latent measure against age. Each latent has been deconfounded from imaging parameters as described in **Supplementary Methods**. Linear model estimates reported in **Supplementary Tables 8 and 9**.

#### Supplementary Figure 9

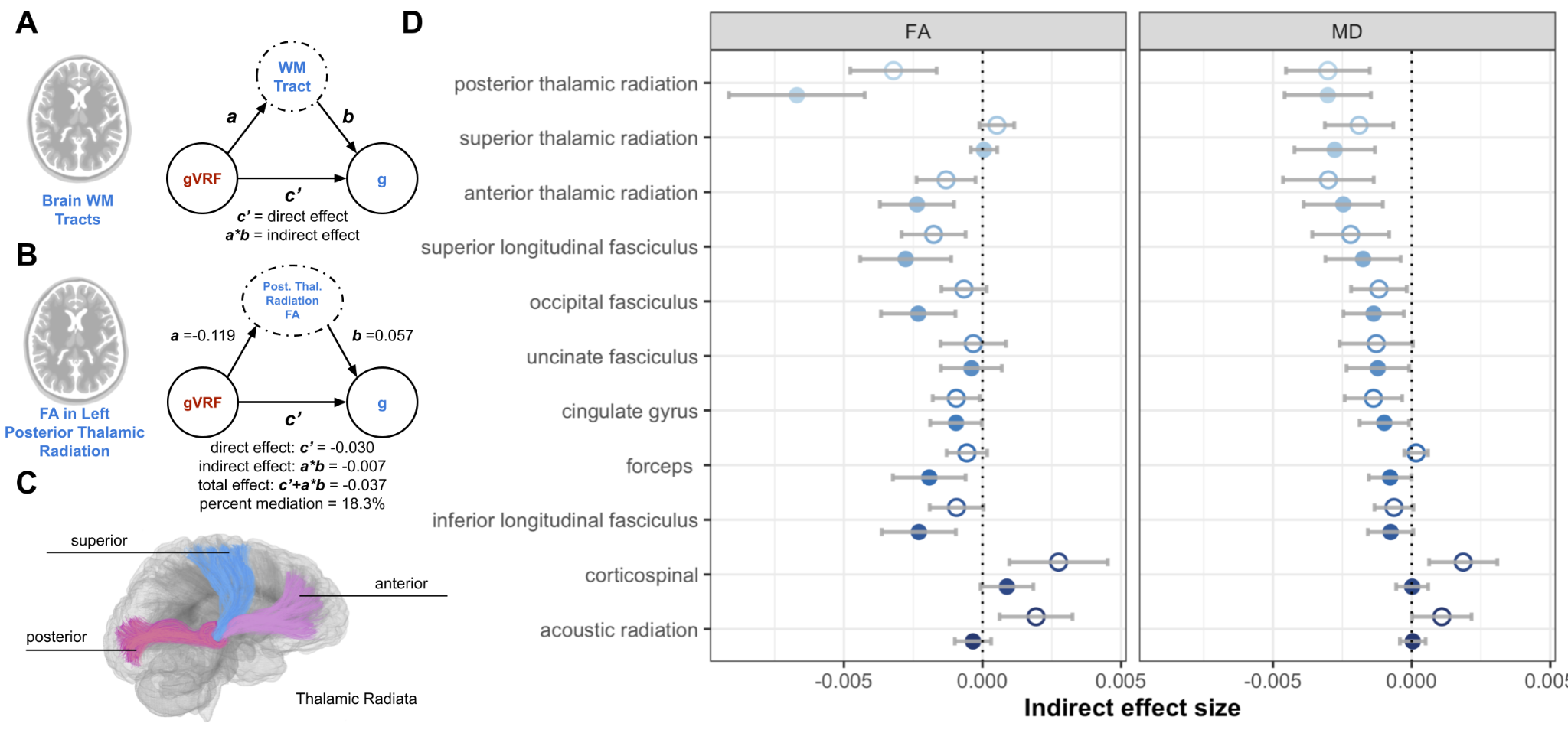

Supplementary Figure 9: White Matter Tract Mediation Modelling

We focus on serial mediation modelling for individual white matter tract quantifiers. (A) Schematic for the modelling procedure for the brain white matter tract quantifiers. (B) Example mediation model for an individual feature, fractional anisotropy of the posterior thalamic radiation. Confidence intervals reported in **Supplementary Table 25**. (C) Illustration of the key white matter tracts with large indirect effects for both FA and MD. (D) Indirect effects for all tested FA and MD measures.

### Supplementary Table Captions

Supplementary Table 1: Summary statistics of the final cohort

Supplementary Table 2: Loadings and Fit Metrics for CFA of Vascular Risk Factors

Supplementary Table 3: Loadings and Fit Metrics for CFA of Cognitive Exams

Supplementary Table 4: PCA Loadings for CMR Radiomics

Supplementary Table 5: Loadings and Fit Metrics for CFA of Fractional Anisotropy and Mean Diffusivity

Supplementary Table 6: PCA Loadings for all Brain MRI IDPs

Supplementary Table 7: CCA Loadings for CMR Radiomics and Brain MRI IDPs

Supplementary Table 8: Association of each Latent Factor with Age and Sex, Raw

Supplementary Table 9: Association of each Latent Factor with Age and Sex, Deconfounded

Supplementary Table 10: Coefficients and Metrics for Pairwise Linear Modelling of all Latents, Raw

Supplementary Table 11: Coefficients and Metrics for Pairwise Linear Modelling of all Latents, Deconfounded

Supplementary Table 12: Propensity Score Matching t-tests

Supplementary Table 13: Single Mediation Model Coefficients and Metrics for all Latents, Raw, lhs: left hand side, op: operation, rhs: right hand side, est.std: standardised estimate

Supplementary Table 14: Single Mediation Model Coefficients and Metrics for all Latents, Deconfounded

Supplementary Table 15: Association of BMI with Heart PC2 controlling for heart size, mediation of the BMI-VNR association by Heart PC2, and mediation of the gVRF-g association by Heart PC2 covarying for BMI

Supplementary Table 16: Multiple Parallel Mediation Model Coefficients and Metrics for all Latents, Raw

Supplementary Table 17: Multiple Parallel Mediation Model Coefficients and Metrics for all Latents, Deconfounded

Supplementary Table 18: Multiple Sequential Mediation Model Coefficients and Metrics for all Latents, Raw

Supplementary Table 19: Multiple Sequential Mediation Model Coefficients and Metrics for all Latents, Deconfounded

Supplementary Table 20: Single Mediation Model Coefficients and Metrics for all Latents and all significant VRF-cognitive pairs, Raw

Supplementary Table 21: Single Mediation Model Coefficients and Metrics for all Latents and all significant VRF-cognitive pairs, Deconfounded

Supplementary Table 22: Single Mediation Model Coefficients and Metrics for all individual CMR radiomics features, Raw

Supplementary Table 23: Single Mediation Model Coefficients and Metrics for all individual CMR radiomics features, Deconfounded

Supplementary Table 24: Single Mediation Model Coefficients and Metrics for all individual BMI IDPs, Raw

Supplementary Table 25: Single Mediation Model Coefficients and Metrics for all individual BMI IDPs, Deconfounded
